# Effectiveness of interventions for modal shift to walking and bike riding: a systematic review with meta-analysis

**DOI:** 10.1101/2024.07.29.24311197

**Authors:** Lauren Pearson, Matthew J Page, Robyn Gerhard, Nyssa Clarke, Meghan Winters, Adrian Bauman, Laolu Arogundade, Ben Beck

## Abstract

**Objective:** To assess the efficacy of interventions aimed at increasing walking and cycling.

**Design:** Systematic review with meta-analysis

**Study selection:** The electronic databases MEDLINE, PsycINFO and Web of Science were searched from inception on 22^nd^ May 2023. Eligible study designs included randomised and non-randomised studies of interventions with specific study design features that enabled estimation of causality. No restrictions on type of outcome measurement, publication date or population age were applied.

**Data extraction and synthesis:** Two reviewers independently extracted data and conducted quality assessment with Joanna Briggs Quality Assessment tools. Studies were categorised by intervention types described within the Behaviour Change Wheel. Where possible, random-effects meta-analyses were used to synthesise results within intervention types.

**Main outcome measures:** The main outcome of interest was modal shift to active modes (walking and cycling). Other outcomes of interest included cycling and walking duration, frequency and counts, active transport duration and frequency, and moderate to vigorous physical activity duration (MVPA).

**Results:** 106 studies that assessed the impact of an intervention on walking, cycling or active transport overall were included. Findings demonstrate that physical environmental restructure interventions, such as protected bike lanes and traffic calming infrastructure, were effective in increasing cycling duration (OR = 1.70, 95% CI 1.20 – 2.22; 2 studies). Other intervention types, including individually tailored behavioural programmes, and provision of e-bikes were also effective for increasing cycling frequency (OR = 1.33, 95% CI 1.23-1.43; 1 study) and duration (OR = 1.13, 95% CI 1.02.-1.22, 1 study). An intensive education programme intervention demonstrated a positive impact on walking duration (OR = 1.96, 95% CI 1.68 – 2.21; 1 study). An individually tailored behavioural programme (OR = 1.23, 95% CI 1.08 – 1.40; 3 studies) and community walking programme (OR = 1.15, 95% CI 1.14 – 1.17; 1 study) also increased the odds of increased walking duration. This body of research would benefit from more rigour in study design to limit lower quality evidence with the potential for bias.

**Conclusions:** This review provides evidence for investment in high-quality active transportation infrastructure, such as protected bike lanes, to improve cycling and active transport participation overall. It also provides evidence for investment in other non-infrastructure interventions. Further research to understand which combinations of intervention types are most effective for modal shift are needed. Active transport research needs to include more robust trials and evaluations with consistent outcome measures to improve quality of evidence and provide evidence on which interventions (or combinations of interventions) are most effective.

**Study registration:** PROSPERO CRD42023445982

**Funding:** This research was funded through the British Columbia Centre for Disease Control, Canada. The research funders did not contribute to the research process or interpretation of findings. The researchers were independent from the funders. Lauren Pearson receives salary support from the National Health and Medical Research Council (GNT2020155). Ben Beck receives an Australian Research Council Future Fellowship (FT210100183).

## INTRODUCTION

The physical and mental health, environmental, social and economic benefits of shifting from private car travel to active travel (walking and bike riding) are well established.^(1–5)^ People who choose active modes of travel are more likely to meet physical activity guidelines,^(2)^ are at lower risk of cardiovascular disease, cancer and mortality,^(2)^ and have higher psychological wellbeing.^(3)^ Shifting trips away from private motor vehicle travel to active travel also acts to reduce vehicular emissions and noise pollution, with positive flow-on health outcomes.^(6)^ Engaging in active travel can also foster social cohesion and community participation.^(7)^

As the world grapples with the critical health challenges of rising physical inactivity and mental health challenges,^(8)^ as well as the escalating impacts of climate change,^(9)^ there is growing recognition of the need to shift travel from private motor vehicles to active travel (e.g. walking and bike riding). Reflecting this, many levels of government in countries around the world are considering investment in modal shift to active travel, including setting modal shift targets.^(10–12)^

Despite the many benefits of walking and bike riding, and increasing recognition and investment from governments, synthesised evidence on the most effective interventions to support modal shift to active travel is lacking. As a result, it is unclear which interventions should be prioritised to maximise uptake of active travel.^(13)^ Identification of priority interventions is further challenged by the myriad interventions available. Examples include providing safe, connected and accessible walking and cycling infrastructure, education campaigns to increase knowledge or understanding of benefits of active travel, mass media campaigns, financial incentivisation, land use policies that enhance access to destinations and services, and policies that restrict car use.^(14, 15)^ Previous systematic and scoping reviews have explored the effects of specific types of interventions to increase participation rates, including education initiatives,^(16)^ infrastructural changes,^(17, 18)^ and environmental changes,^(19)^ or have grouped interventions broadly as ‘carrots’ or ‘sticks’.^(20, 21)^ The limitations of these earlier reviews include a focus on single specific intervention types which prevents comparison across different intervention types, variation in outcomes, and given the year of publication, they are missing the growing body of evidence. As a result, there is a lack of an up-to-date synthesis of evidence on the effectiveness of interventions that promote modal shift to walking and/or bike riding. Therefore, this systematic review and meta-analysis aims to measure the effectiveness of interventions to increase active travel with a primary outcome of modal shift.

## METHODS

The protocol for this systematic review and meta-analysis was registered with PROSPERO (CRD42023445982). We have reported this paper in accordance with the Preferred Reporting Items for Systematic reviews and Meta-Analyses (PRISMA) 2020 Guidelines (Data Supplement 1).^(22)^

### Eligibility criteria

We included studies that enrolled participants of any age, including children, adolescents and adults. We included all intervention types that incentivised or motivated people to use active transport, rather than used punishment to induce this behaviour. The primary outcomes of interest were modal shift measured through mode frequency or duration. Modal shift refers to a change in transport mode, from one mode (such as driving a motor vehicle) to another (such as bike riding or walking) for a particular trip or journey, or in terms of overall long-term travel behaviour. Frequency refers to the number of trips taken by a particular mode, while duration refers to the time taken on that mode. Secondary outcomes included: changes in the frequency of trips by walking and/or bike riding, and time spent walking and/or bike riding.

We included randomised trials and non-randomised studies of interventions, including controlled before-after studies and interrupted time series studies. Explicit study design features were used to categorise study design, rather than how authors described the study design within each article. This was due to study design labels being used inconsistently across some non-randomised studies. A checklist of study design features was used during the selection process to ensure inclusion of eligible/appropriate non-randomised studies.^(23)^

### Exclusion criteria

We excluded studies that exclusively sampled competitive and/or sports cyclists and studies that focussed on enhancing performance in sport cycling. Given the focus of this review on modal shift, we excluded studies that focused on walking and bike riding specifically for recreational purposes. Recreational trips include those made for leisure or exercise purposes, rather than those trips with an intent to reach a destination; there are differences in the factors that affect trips for recreation as opposed to trips for transport purposes (such as commuting to work, travelling to school and travelling to shops).^(24)^ Given the focus on positive reinforcement interventions, interventions that involved restriction or coercion were excluded.

### Search and selection methods

We searched peer-reviewed articles indexed within Ovid Medline, Ovid PsycINFO and Web of Science to identity articles published from inception up to 22^nd^ of May 2023, and reviewed the reference list of relevant systematic reviews to identify additional eligible studies. The search strategies are shown in Data Supplement 2. No language restrictions were applied to the search, however only articles published in English were included in the review. No restrictions were placed on date of publication. We included studies if they measured the impact of an intervention aimed to increase walking, cycling or active transport overall, regardless of the type of outcome measurement, as long as they met all other eligibility criteria listed above.

Two authors independently screened titles and abstracts of all records yielded by the search, and all full-text reports deemed (potentially) eligible. Any disagreements were firstly resolved through discussion, or if required, through consultation with a third author who acted as the adjudicator.

### Data extraction

Two authors independently extracted data from each included study using a data extraction form developed specifically for this review. Authors resolved any discrepancies in extraction through discussion or adjudication from a third author. Intervention types were categorised within the Kelly *et al.*^(14)^ classification system, adapted from the Behaviour Change Wheel.^(25)^ Where interventions were classified with more than one category, two authors assigned a main category and the remaining categories were assigned as subclassifications. The main intervention type was decided based on the criteria listed in Kelly *et al*.^10^, and based on the main aim of the intervention. Categories included:

- Education interventions (aimed at increasing knowledge)
- Persuasive interventions including campaigns (using communication to stimulate action)
- Incentivisation (creating expectation of reward)
- Training (imparting skills)
- Environmental restructuring (change of physical and social environments)
- Modelling interventions (buddying systems and mentoring)
- Enablement interventions (provision of resources)

### Quality assessment

The Joanna Briggs Institute Critical Appraisal Tools were used to assess the methodological rigour of included studies.^(26)^ The tools are specific to study design and include closed questions regarding the methodology, recruitment, analysis and conclusions of each study. Two authors independently conducted quality assessments for each included study. Results of quality assessments were compared between authors. If differences of judgements arose, the two assessors discussed these until consensus was reached, or consulted with a third author where necessary.

### Meta-analysis

Two types of outcome data were extracted for this review, including number of trips within a defined time period made by active modes, and mean number of trips within a defined time period, with measures of variability (e.g. standard deviations). To be included in a meta-analysis, two or more studies were required to have similar study populations, interventions and follow-up time periods, and identical measured used (for example, number of trips taken by bike). Studies were grouped by intervention and outcome type (for example, the impact of educational interventions on cycling duration), and synthesised via random-effects meta-analyses conducted in RStudio with the “meta” and “metafor” packages.^(27, 28)^ The inverse-variance method was used to weight studies, the DerSimonian and Laird^(29)^ method of moments estimator was used to estimate the between-study variance, and the 95% confidence intervals (CIs) for the summary estimates were calculated using the Wald type method. Heterogeneity was assessed visually by inspecting the forest plots and quantitatively with the I^2^ statistic to measure inconsistency.^(30)^ To enable comparison of results of meta-analyses, those that produced a difference in means or in frequency of outcome were transformed to an odds ratio following methods outlined in the Cochrane Handbook.^(31)^ For studies that reported a mean and standard deviation for each group, we first generated a standardised mean difference (SMD) via the Cohen’s d formula. Following this, the SMD was transformed into a log odds ratio via the following formula:

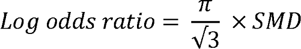

The log odds ratio was exponentiated to become an odds ratio. When frequency of an outcome was reported alongside the participant sample size, log odds ratio could be calculated using the ‘metafor’ package^(28)^ within RStudio, and exponentiated to produce an odds ratio.

## RESULTS

Our search identified 21,550 records for review (Figure 1). After removal of duplicates, we screened 9,500 records against selection criteria, and retrieved 217 references for full text screening. One report was not able to be retrieved as it was published in a journal written in a language other than English.^(32)^ We excluded 111 studies that did not meet inclusion criteria. The majority of studies were excluded as they did not have a control group (*n*=65), or did not measure the outcomes of interest (*n*=23). A total of 106 reports of 104 studies were included within the final review.

**Figure 1.**
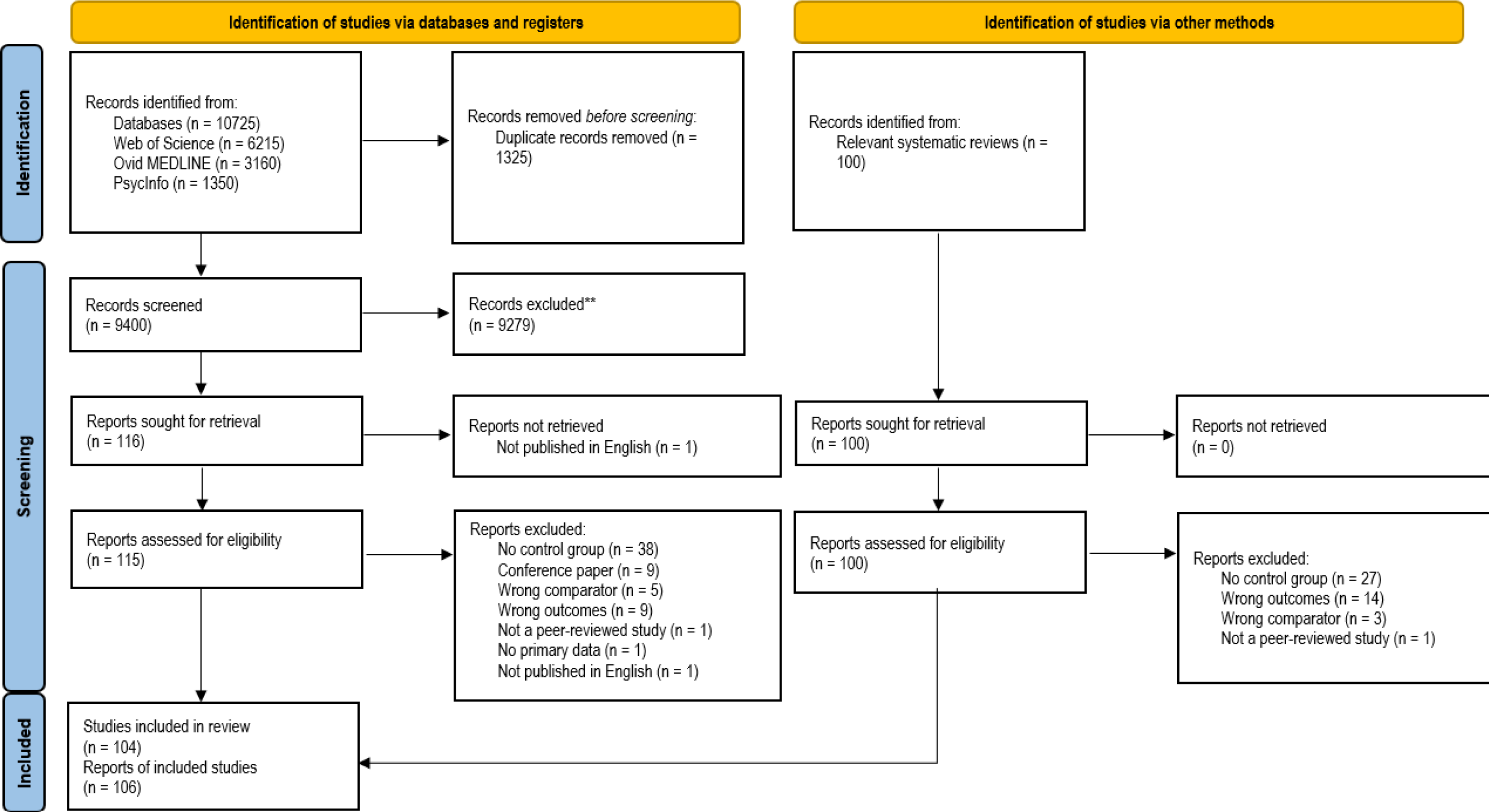
PRISMA flow diagram of search result.

### Study Characteristics

#### Study Design, Population and Setting

The most common study designs were controlled before-after studies (*n*=68, 64%) followed by randomised controlled trials (*n*=36, 34%), the least common was interrupted time series, used by 2% (*n*=2) of included reports. The studies were conducted in 23 different countries, the most common of which were the USA (*n*=32), Australia (*n*=14), England (*n*=9), Canada (*n*=5) and China (*n*=5). Studies were published between 1983 and 2023. Forty-three studies (41%) were conducted in schools and eight in workplaces (8%). The participants of studies included in this review were aged between 5 and 79 years, and sample sizes ranged from 11 to 26,231. The proportion of women in the intervention groups ranged between 12 to 100%, with a median of 52%. Twenty-three studies did not report the proportion by gender and/or sex within the sample. A detailed description of each study population can be found in Data Supplement 3, and summary of characteristics in Table 1.

**Table 1.**
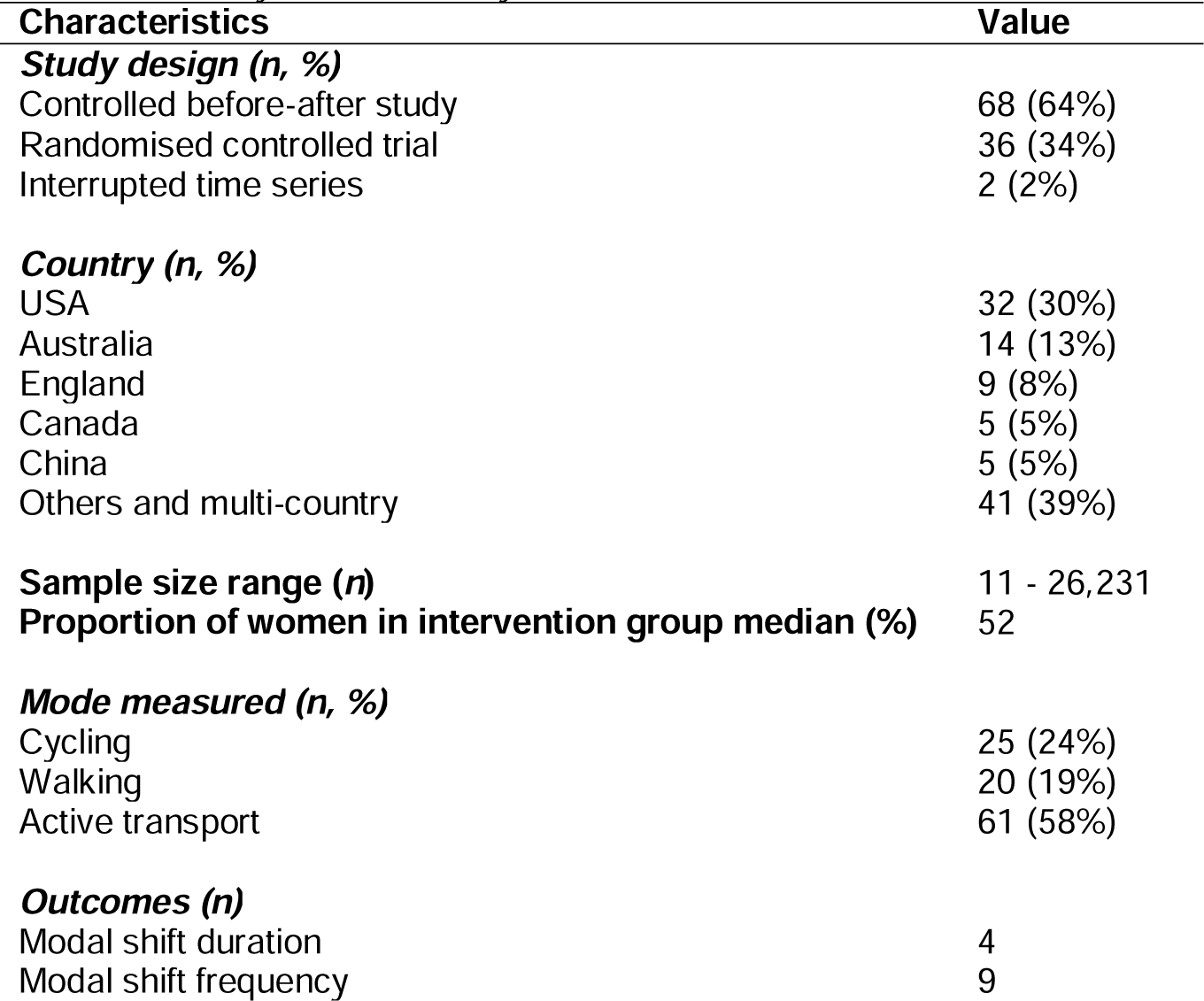
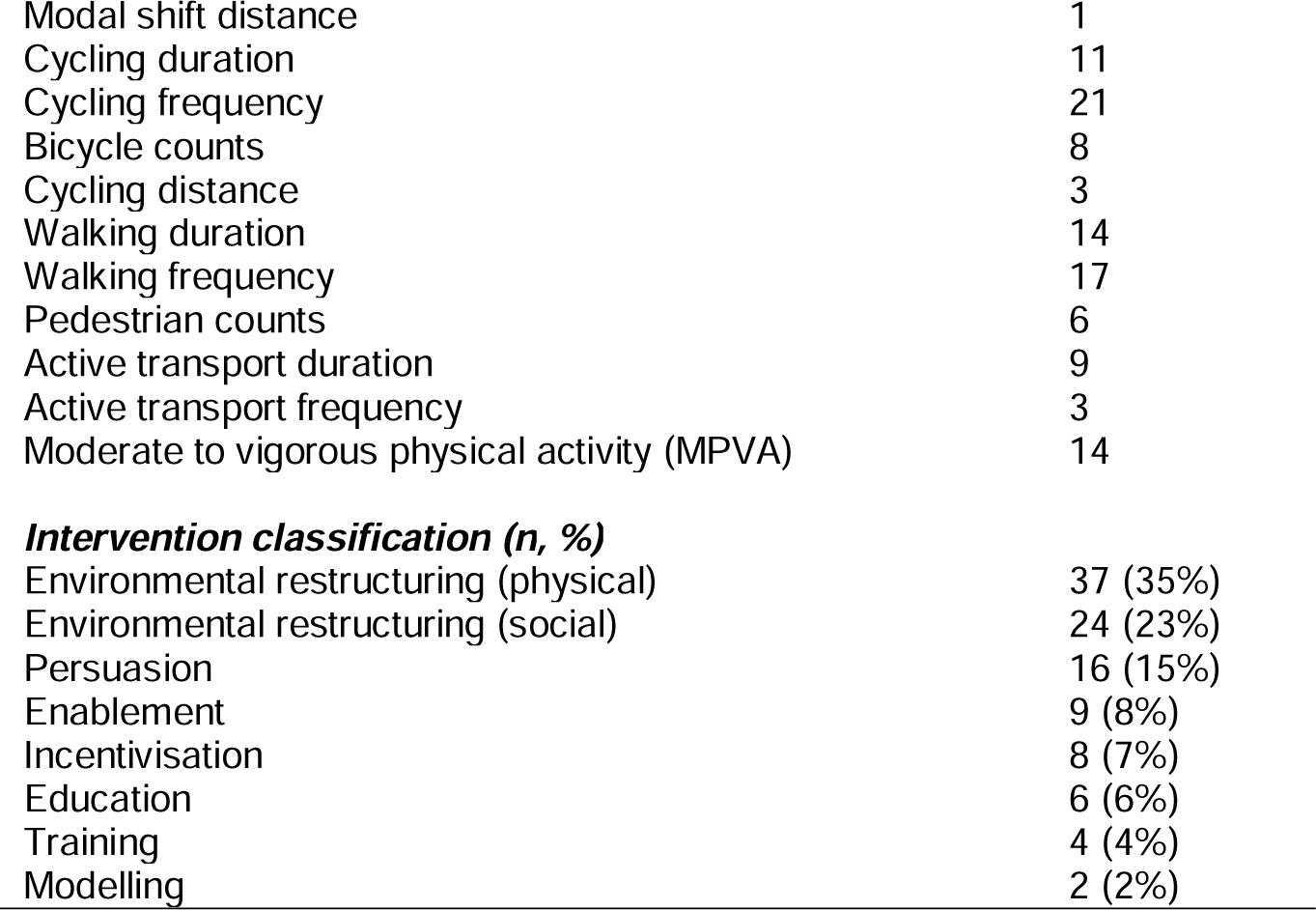
Summary of included study characteristics.

| Characteristics | Value |
| --- | --- |
| **Study design (n, %)** |  |
| Controlled before-after study | 68 (64%) |
| Randomised controlled trial | 36 (34%) |
| Interrupted time series | 2 (2%) |
| **Country (n, %)** |  |
| USA | 32 (30%) |
| Australia | 14 (13%) |
| England | 9 (8%) |
| Canada | 5 (5%) |
| China | 5 (5%) |
| Others and multi-country | 41 (39%) |
| **Sample size range (n)** | 11 - 26,231 |
| **Proportion of women in intervention group median (%)** | 52 |
| **Mode measured (n, %)** |  |
| Cycling | 25 (24%) |
| Walking | 20 (19%) |
| Active transport | 61 (58%) |
| **Outcomes (n)** |  |
| Modal shift duration | 4 |
| Modal shift frequency | 9 |
| Modal shift distance | 1 |
| Cycling duration | 11 |
| Cycling frequency | 21 |
| Bicycle counts | 8 |
| Cycling distance | 3 |
| Walking duration | 14 |
| Walking frequency | 17 |
| Pedestrian counts | 6 |
| Active transport duration | 9 |
| Active transport frequency | 3 |
| Moderate to vigorous physical activity (MPVA) | 14 |
| ***Intervention classification (n, %)*** |  |
| Environmental restructuring (physical) | 37 (35%) |
| Environmental restructuring (social) | 24 (23%) |
| Persuasion | 16 (15%) |
| Enablement | 9 (8%) |
| Incentivisation | 8 (7%) |
| Education | 6 (6%) |
| Training | 4 (4%) |
| Modelling | 2 (2%) |

#### Outcomes

The majority of the studies included in this review focused on the effects of interventions on active transport overall (55%, *n*=58) with no disaggregation by active transport mode. Twenty-five studies (24%) focused on cycling only, twenty (19%) focused on walking only and three studies reported combinations of walking, cycling and active transport. Only 14 studies specifically measured the primary outcome of interest (modal shift) either through duration of use (*n*=4), frequency of use (*n*=9) or trip distance (*n*=1). Other outcomes identified in this review included duration, frequency, distance, count, moderate-to-vigorous physical activity (MVPA), each within either walking, cycling or active transport overall. Further details of outcomes included are described in Table 1.

#### Interventions

Most studies (*n*=65) were classified under a single main intervention type, of which the most common was physical environment restructuring (*n*=37) and social environmental restructuring (*n*=24). Forty-one of the included studies had both a main intervention type classification and intervention type subclassifications, such as those that delivered infrastructural interventions (physical environmental restructure) interventions alongside wider social campaigns (social environmental restructure). Further details of interventions provided are provided in Data Supplement 3.

### Quality Assessment

Quality assessments were done on the 32 randomised controlled trial studies (see Data Supplement 4). Assessment revealed limited reporting of blinding of participants, assessors, and individuals delivering the intervention. No studies reported blinding of those delivering the intervention to the group participants were randomised to, and one study reported information relating to participants being blinded to which treatment group they were assigned to. This is likely due to the difficulties and feasibility in blinding in transport intervention research. Similarly, four studies reported blinding assessors to which group participants were randomised to ^(33–36)^. Outcome measurement was reliable across all studies, including consistent measurement across treatment groups.

Studies not classified as randomised controlled trials were assessed for quality using tools specific to quasi-experimental studies (see Data Supplement 4). Notable areas of concern included intervention and control groups not being similar in characteristics at baseline, and either follow-up measures not being taken or differences in follow up not described and analysed. Thirty-nine studies did not report intervention and control groups that were appropriately similar at baseline. This was due either through not reporting characteristics of intervention and control groups, or through having groups with statistically significant difference in characteristics. Further details for assessed studies are outlined in Supplementary Material.

### Modal Shift Outcomes

#### Modal Shift Duration

Modal shift duration was used to measure intervention effectiveness in four studies ^(37–40)^. Increased time spent bike riding and walking was observed in three studies, which delivered an educational intervention, a physical environmental restructuring intervention and a social environmental restructuring intervention. Two studies observed a reduction in car or motorised vehicle travel, including a 7.7 minutes per week reduction following implementation of supportive active transport infrastructure^(37, 38)^, and 3.08 minutes per week reduction from general health messaging.^41^ We could not calculate 95% CIs for any of the mean differences as no measures of variability were available. Thus there is uncertainty in the effects of these interventions on modal shift duration.

**Table 2.**
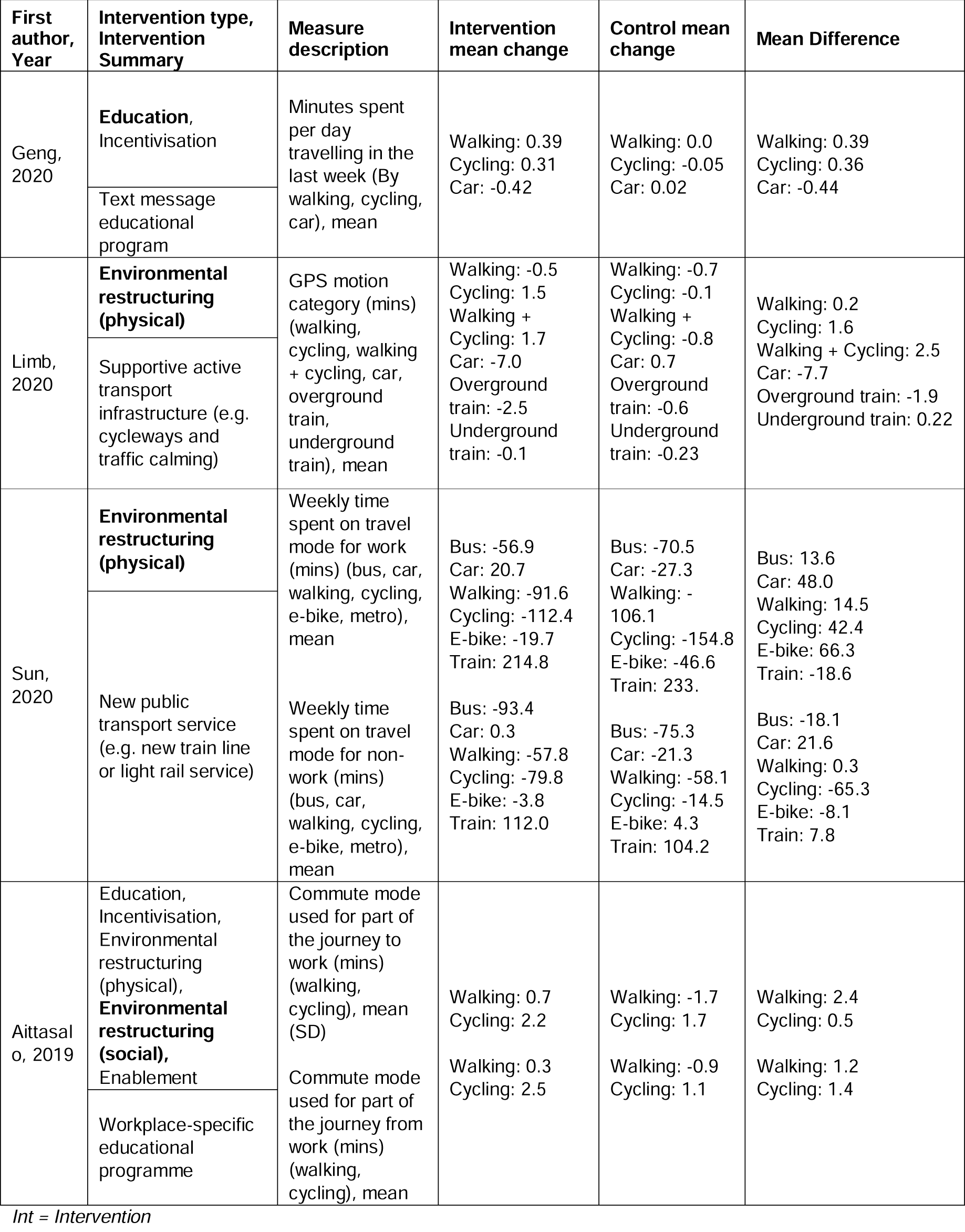
Effect of interventions on modal shift duration.

#### Modal Shift Frequency

Sixteen studies used the number of trips by travel mode to assess intervention effectiveness on modal shift ^(37, 40–52)^. Five studies examined social environmental interventions, three used educational interventions, three used a physical environmental restructure, three used enablement, one used persuasion and one used incentivisation. A social environmental restructure intervention involving behavioural strategies within a workplace recorded an 11.1% reduction in the proportion of people travelling to work by car or motorcycle, a 4.8% increase in trips by bike and an 8.2% increase in walking trips.^(40)^ Following three months’ access to an e-bike with trailer, longtail bike or traditional bike with trailer, Bjornara *et al.*^(51)^ noted a 4.6% increase in trips to kindergarten made by bike, by participants previously categorised as car-users. We could not calculate 95% CIs for any of the mean differences because a denominator for total trips was not available. There is therefore, uncertainty in the effects of these interventions on modal shift frequency.

**Table 3.**
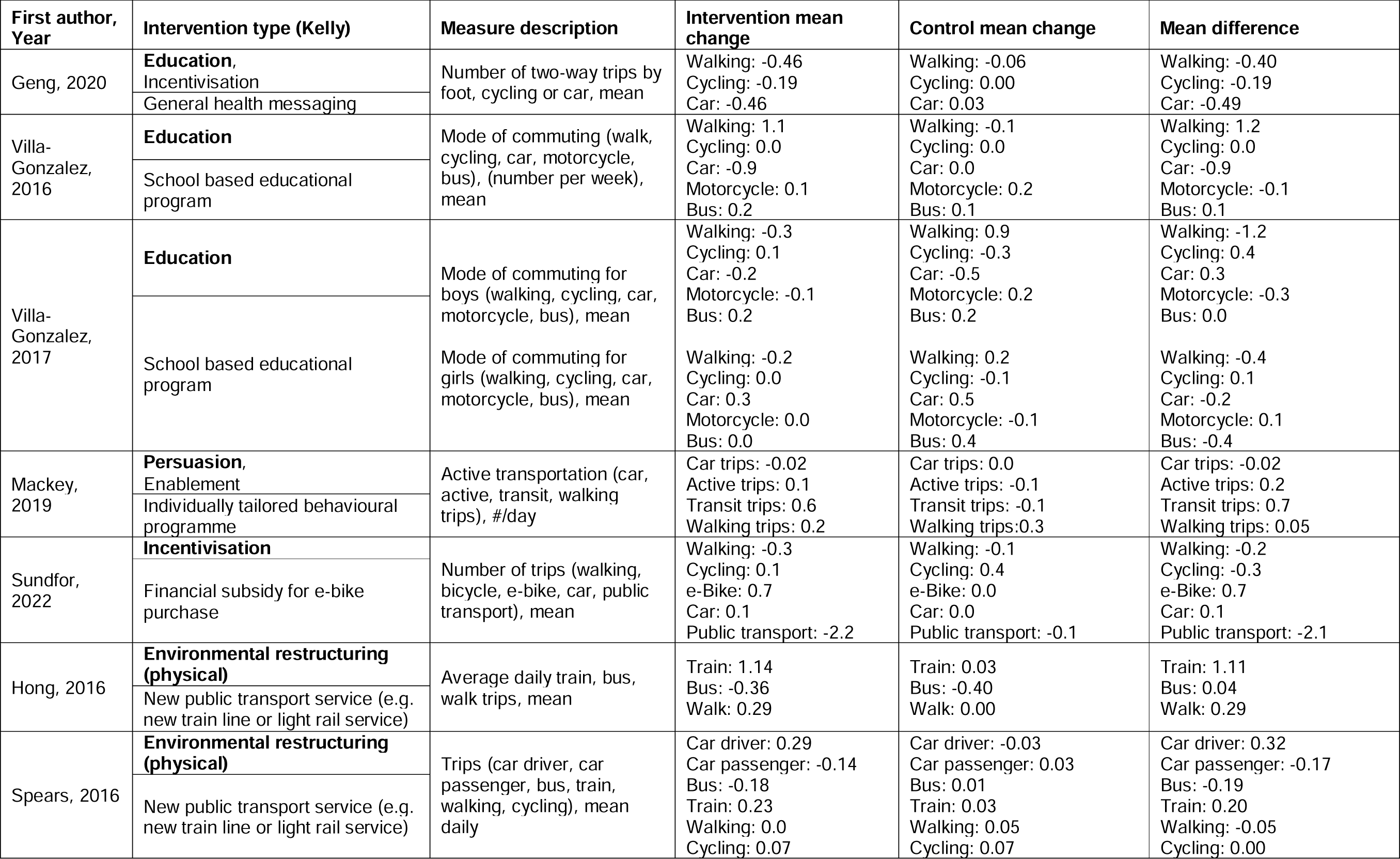
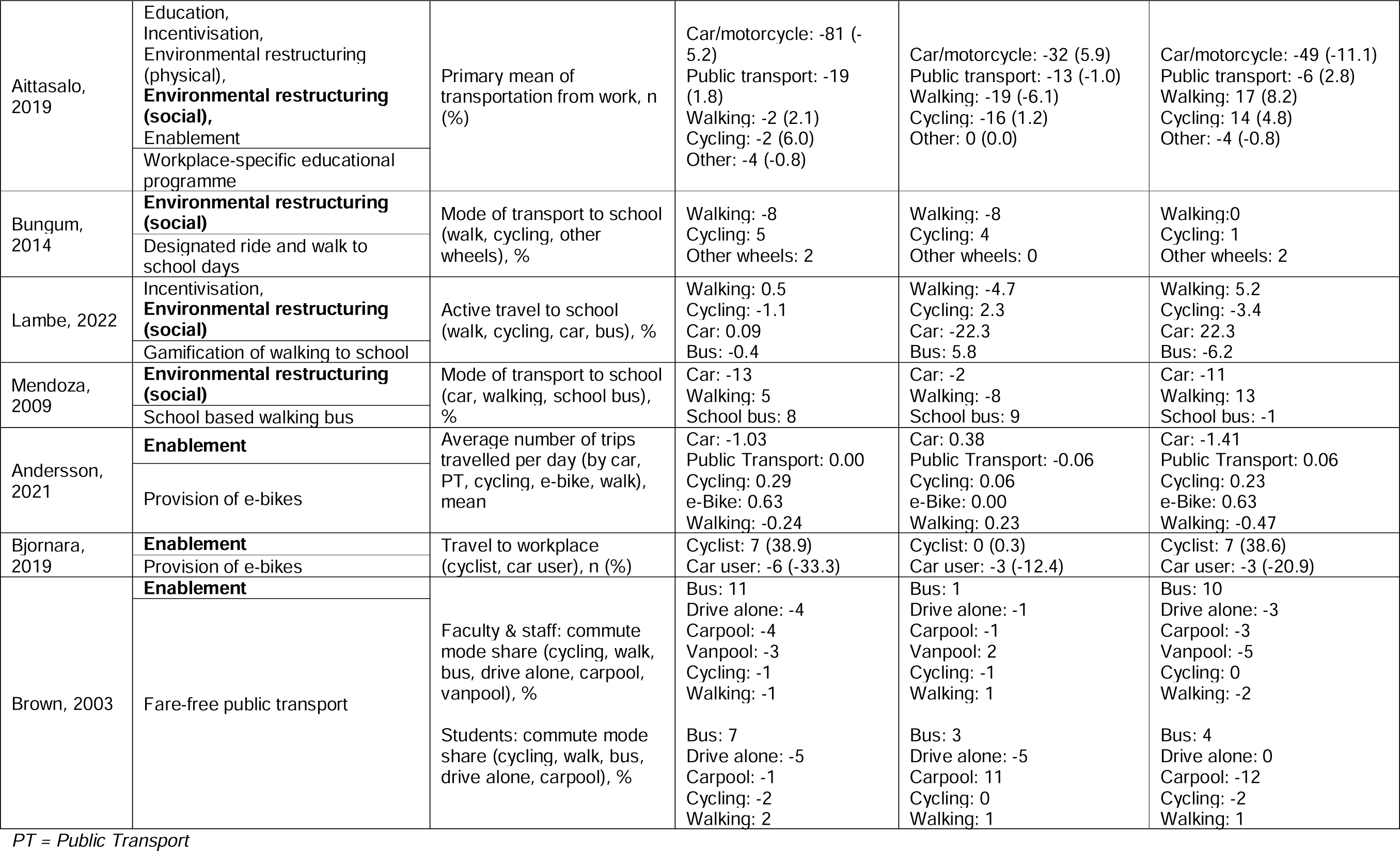
Effect of interventions on modal shift frequency.

### Cycling Outcomes

Eleven studies measured cycling duration and/or frequency and were able to be either meta-analysed (*n*=4) or displayed as single point estimates within Figure 2. Supportive active transport infrastructure (e.g. cycleways and/or traffic calming infrastructure) had the greatest impact on cycling duration compared to control (OR = 1.70, 95% CI 1.20 – 2.22; 2 studies, 1424 participants). Educational interventions (specifically an intensive education programme) had the next greatest impact on cycling duration (OR = 1.33, 95% CI 1.23-1.43; 1 study, 620 participants), and provision of e-bikes had the greatest impact on cycling frequency (OR = 1.13, 95% CI 1.02.-1.22; 1 study, 65 participants). All other studies included had 95% CIs crossing the null.

**Figure 2.**
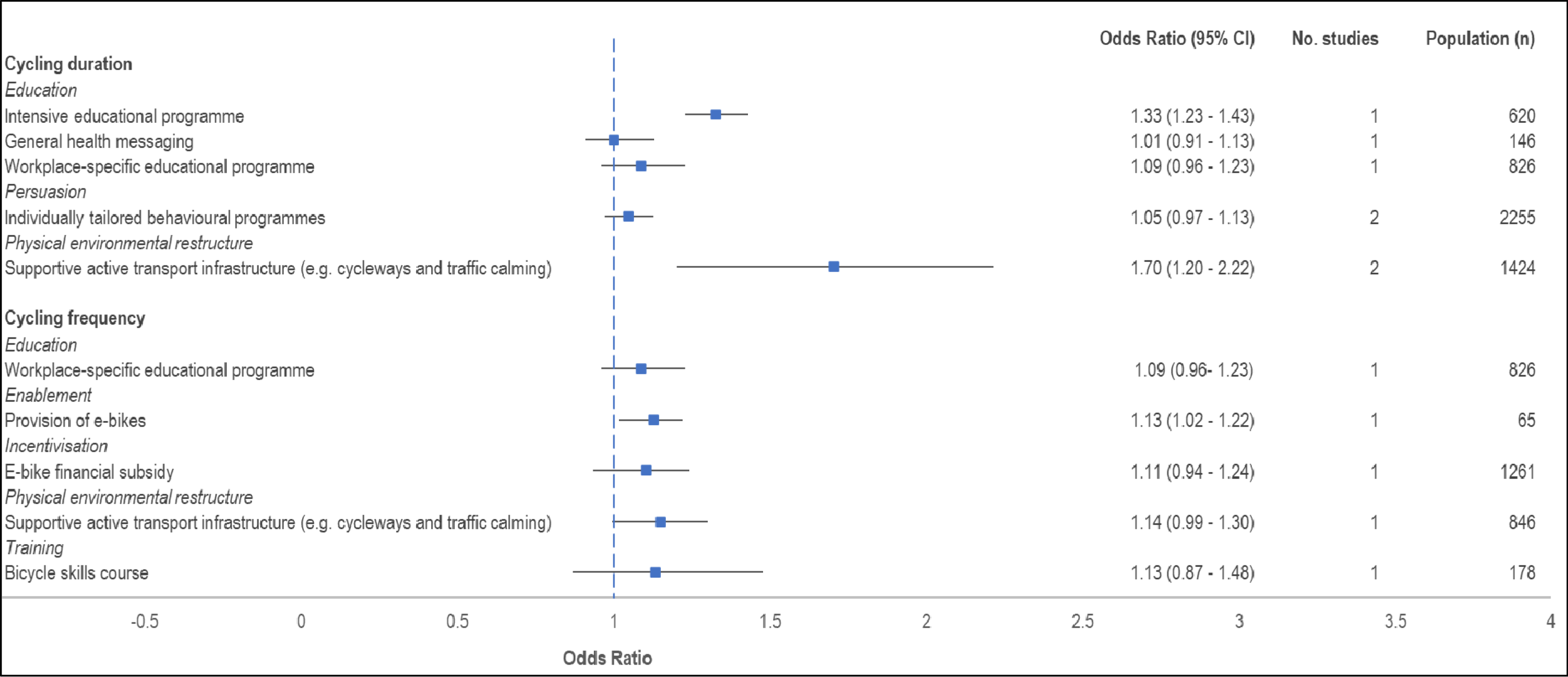
Forest plot of meta-analyses and single point estimates for cycling duration and frequency outcomes.

#### Cycling Duration

Of the 11 studies that measured cycling duration, 4 were meta-analysed within intervention classifications (physical environmental restructuring and persuasion) ^(34, 37, 38, 40, 53–60)^. Individual mean differences are reported in Table 4. Three educational interventions, including an intensive educational programme^(53)^, and a campaign of general health messaging^(37)^ and workplace specific educational programme measured cycling duration^(40, 59)^, however demonstrated substantially different results. The intensive 16-week program of education sessions (Salinas *et al.*) increased weekly cycling duration by 14 minutes (MD = 14, 95% CI 13.1 – 14.1). In comparison, in a workplace-specific education program involving behavioural seminars, Aittasalo *et al.* reported a 1.4-minute increase in weekly cycling, which crossed the null (MD = 1.4, 95% CI -0.7 – 3.5). Results of individual studies are presented in Data Supplement 5.

Meta-analysis of two studies evaluating persuasion interventions (see Figure 3) and their impact on weekly cycling minutes demonstrated a mean difference in favour of the intervention of 5.01 minutes (95% CI -3.08 – 13.10; I^2^ 0%); although the 95% CI crossed the null. Studies included within the meta-analysis included an electronic health-focused intervention focused on self-regulation^(54)^ and a series of information for increasing physical activity tailored to individuals’ surrounding environments.^(55)^

**Figure 3.**
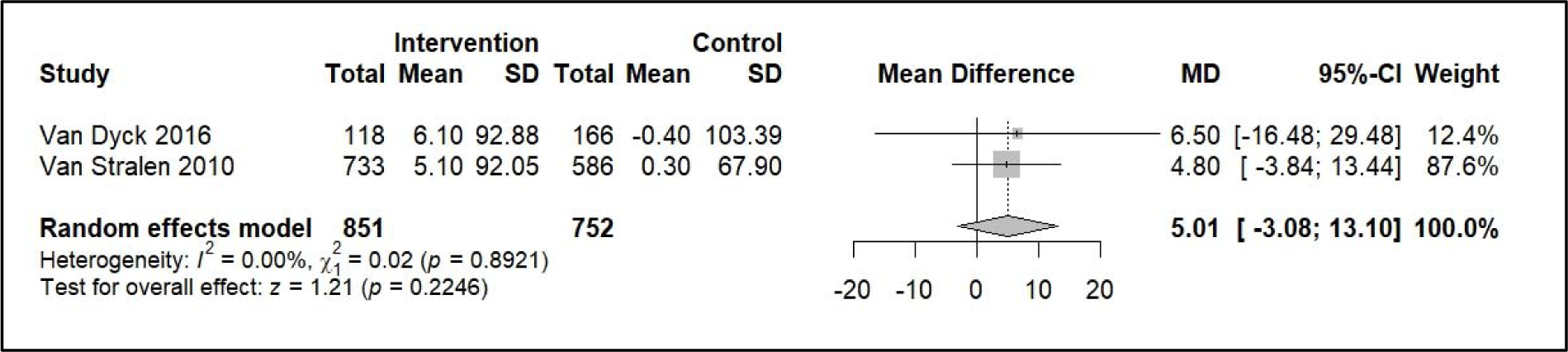
Meta-analysis of persuasion interventions on cycling minutes weekly.

Meta-analysis of two studies evaluating physical environmental restructure interventions (see Figure 4) demonstrated increased cycling minutes per week of 28.7 minutes higher in the intervention group compared with the control (95% CI 18.6 – 38.8; I^2^ 0%). Both studies within this meta-analysis delivered protected cycleways^(57)^ however Limb *et al.* also involved implementation of a purpose-built mixed-use residential development based on active living design principles.^(38)^

**Figure 4.**
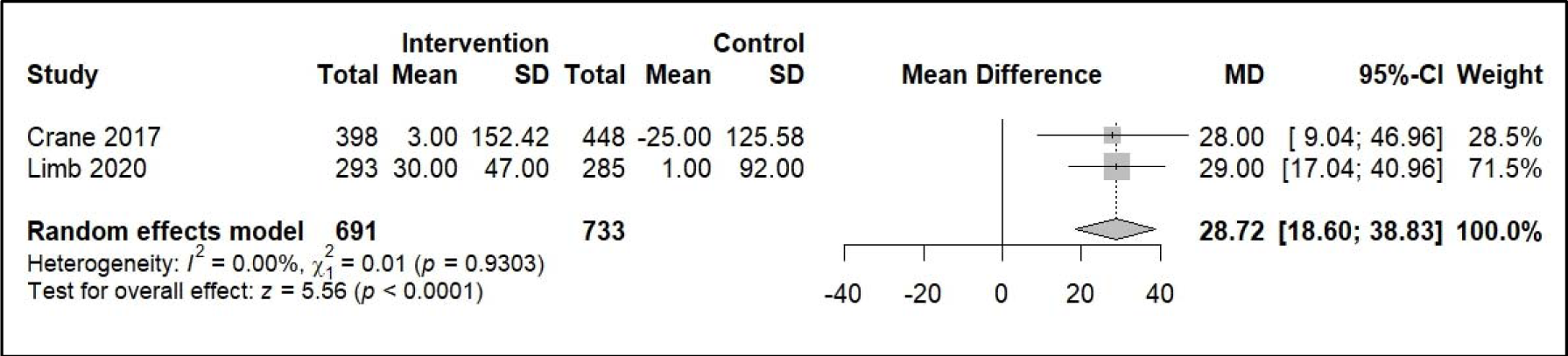
Meta-analysis of physical environmental restructure interventions on cycling minutes weekly.

#### Cycling Frequency

Twenty studies included in the review measured cycling frequency ^(40–42, 46–48, 50–52, 57, 58, 60–68)^. There was substantial variation in the measurement of cycling frequency, leading to no studies being suitable for meta-analysis. Studies that measured cycling frequency reported this as either a proportion (for example, proportion of people cycling weekly) or as a mean (for example, average number of days cycling per week). Results of individual studies are presented in Data Supplement 5.

Several of the studies which used cycling frequency as a measure were conducted in school settings. An enablement intervention involving a daily “bicycle train”, where students cycled with staff to and from school demonstrated a 42% increase in daily cycling to school.^(63)^ Bungum *et al.*^(47)^ implemented a “Nevada Moves Day” event, encouraging students to walk or bicycle to school, demonstrating no increase in students who reported using a bicycle as their main mode of transport to school.

In a workplace setting, Bjornara et al. demonstrated a 38.6% increase in the proportion of people cycling to work following provision of bikes and e-bikes. A 95% CI could not be calculated as measures of variance were not available. There is therefore, uncertainty in the effect of this intervention on cycling frequency.

### Walking Outcomes

Thirteen studies measured walking duration and/or frequency and were able to be either meta-analysed (*n*=3) or displayed as single point estimates within Figure 3. One intensive education programme intervention demonstrated the greatest increase in odds of increased walking duration when compared with control (OR = 1.96, 95% CI 1.68 – 2.21; 1 study, 285 participants). Individually tailored behavioural programmes (OR = 1.23, 95% CI 1.08 – 1.40; 3 studies, 749 participants) and community walking programmes (OR = 1.15, 95% CI 1.14 – 1.17; 1 study, 573 participants) also increased the odds of increased walking duration. All other studies included as single point estimates showed a reduction in the odds of increased walking duration or frequency, or had 95% CIs crossing the null.

**Figure 3.**
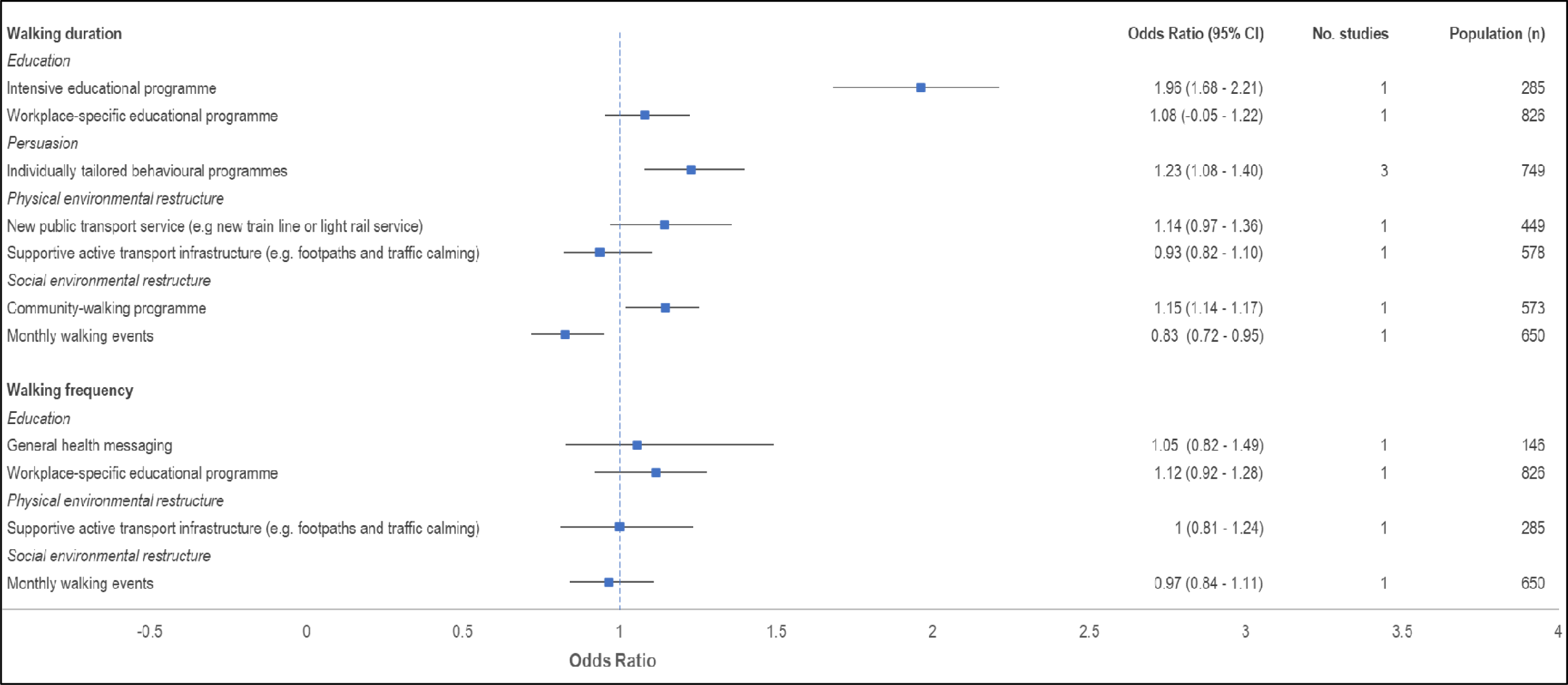
Forest plot of meta-analysis and single point estimates for walking duration and frequency outcomes.

#### Walking Duration

Fourteen studies measured walking duration, three of which were able to be meta-analysed within the persuasion intervention classification ^(36, 38, 40, 53–55, 59, 69–75)^. Meta-analysis of these studies (see Figure 5) involving individually tailored behavioural programmes indicated a 27.2-minute increase in weekly minutes walked following intervention delivery (95% CI 10.1 – 44.3; I^2^ 0%). Across studies, an education intervention involving an intensive educational programme had the greatest impact, increasing walking by 39 minutes per week on average compared with the control (MD = 39 minutes per week; 95% CI 21.1-46.3). Two studies that delivered physical environmental changes demonstrated varied results. He et al.^(73)^ involved implementation of a new train line, resulting in an increase of 48 weekly minutes walked compared to the control (95% CI -42.97 – 138.97). Limb et al. ^(38)^, reported no change in weekly minutes walked following implementation of a purpose-built residential development based on active living principles. Results of individual studies are presented in Data Supplement 5.

**Figure 5.**
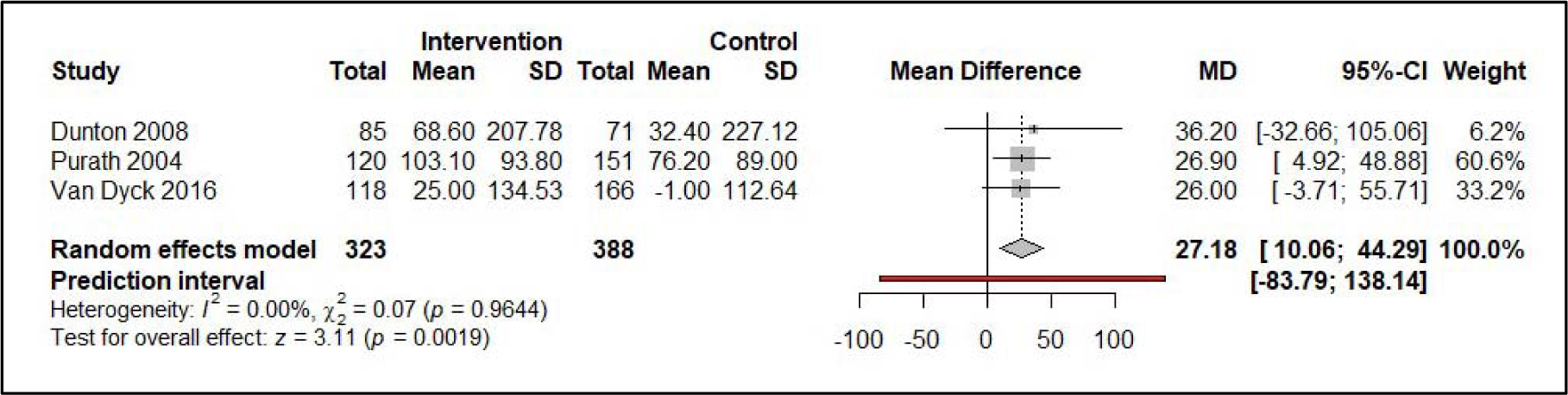
Meta-analysis of persuasion interventions impact on weekly minutes walked.

#### Walking Frequency

Frequency of walking was measured in 17 studies ^(37, 40, 41, 43, 44, 46–50, 59, 75, 77–80)^. Similar to cycling frequency, there was substantial variation in the measurement of walking frequency, and outcomes were either expressed as a proportion or mean. There was variation in the effect estimates displayed in Data Supplement 5, some of which decreased walking frequency. Similar to the cycling frequency results for the same study, Brown et al^(79)^ implemented a free bus to a university campus, resulting in decreased walking to and from campus. Lambe et al^(48)^ implemented a Beat the Street program which increased walking frequency by 36.7% but decreased cycling frequency. Mendoza^(49)^ implemented a walking school bus intervention resulting in an increase of walking frequency of 13%.

## DISCUSSION

### Statement of the principal findings

This systematic review with meta-analysis identified a range of interventions from 106 reports that evaluated uptake of active modes of transport. We demonstrated that physical environmental restructuring interventions including cycleways and traffic calming infrastructure led to the greatest increases in cycling duration (28 minutes per week), and that educational and persuasion interventions involving individually tailored behavioural programmes or intensive workplace-specific programmes were most effective for increasing walking for transport. Within school settings, bicycle bus/train interventions resulted in a 42% increase in bike riding to school, and a walking school bus resulted in a 38% increase in overall active transport. Abundant high-quality evidence for the impact of interventions on modal shift was lacking, limiting our understanding of the effectiveness of interventions in shifting people from sedentary modes of travel (such as by private car) to active modes. Variation in outcome reporting also limited synthesis of findings within intervention types and outcome types. While some results demonstrated promising impacts on active transport, these were almost always based on findings of a single study, and most studies had methodological limitations and were conducted in a variety of different built environment contexts. Further evidence of intervention effectiveness with consistent measures and robust study design is required to draw further conclusions of effectiveness of individual intervention types and understand generalisability of findings.

### Strengths and weakness of the study

Due to heterogeneity in interventions, meta-analysis of like interventions was not possible in the majority of cases, and as a result many point estimates reflected the results of a single study. We were also unable to account for combinations of interventions, such as Brown, 2016^(79)^ which provided both a new public transport service and cycleway, and meta-analysis was reliant on grouping by the main intervention classification (as assessed by the author team). Comparisons through meta-analyses were only possible when the intervention, sample, study design and outcome measure were similar. As a result, the forest plots presented reflect only a subset of studies included in this review. We suggest the reader use both a combination of the meta-analysis results and the tables (reporting individual study results) when interpreting findings. We did not consider interventions to limit driving or interventions to increase the use of public transportation, both of which may serve to shift people towards active transportation. The great majority of included studies were conducted within high-income countries, limiting generalisability to low- and middle-income countries (LMIC).

### Strengths and weaknesses in relation to other studies & implications for policymakers

This review identified that physical environmental restructuring interventions, including infrastructure such as protected cycleways, are effective for increasing cycling duration. Findings relating to the impact of infrastructure on cycling have been reported in previous systematic, literature and policy reviews, many of which included study designs not considered in this review (because of risk of bias associated with these study designs). For example, Karmanieme *et al.*^(81)^ systematically reviewed 51 prospective cohort studies and natural experiments, identifying that cycling paths, traffic free routes and bike lanes predicted increases in cycling frequency. Similarly, Stewart *et al.*^(82)^ reviewed 12 studies which specifically aimed to increase commuter cycling through infrastructural measures, identifying small increases in participation within large populations following implementation of environmental approaches, such as city-wide bicycle infrastructure. It is well established that the biggest barrier to participation in bike riding is how safe someone feels when they are riding a bike, particularly in the presence of motor vehicle traffic^(83)^. Infrastructure that minimises interactions between people cycling and motor vehicle traffic, such as protected cycleways, traffic free routes and low urban speed limits, enhance perceived safety and are of greater preference to less confident riders, women and older adults^(24, 84)^. In order to achieve the wide environmental, health, economic and social benefits that come from greater participation in active transport, implementation of high-quality protected and connected infrastructure that limits interactions between pedestrians, riders and motor vehicles is critical.

While physical environmental restructure interventions were most effective in increasing cycling participation and frequency, other types of interventions showed some evidence of efficacy. This included persuasion interventions, particularly individually tailored behavioural programmes, and education, involving intensive educational programmes. Notably, this review did not consider the effect of combinations of intervention types, and how this compares to the outcomes of sole intervention categories. Winters *et al.*^(85)^ conducted a review of policies that increase active travel, showing that while provision of physical environments conducive to safe and convenient active travel are key to increasing participation, policies are most effective when implemented in combination with other intervention types. These combinations of interventions, delivered as a “comprehensive package” are particularly effective when targeting the different levels of the socio-ecological framework (i.e. individual, interpersonal, community, society and city). In practice, this may involve combining infrastructural changes such as implementation of cycleways (physical environmental restructure) to target city and society-level factors, social environmental restructure interventions to target community and interpersonal factors, and persuasion interventions to target the individual. Further, combinations that integrate interventions to both promote the uptake of active travel, and reduce car use may be more effective for modal shift^(86)^.

School-based interventions formed a substantial proportion (41%) of the studies included within this review. Within these, studies which implemented walking or cycling “school buses” or “trains” where children walked or biked to school within a group with an adult chaperone, demonstrated substantial increases in active transport participation. In a review of outcomes and barriers of walking school buses, Smith *et al.*^(87)^ noted that while these interventions often resulted in positive outcomes, their sustainability was limited, often relying on volunteer programs. Their success can also be dependent on the availability of surrounding infrastructure to support safe use and uptake^(88)^. Despite this, their implementation within communities with limited uptake in active travel, could act as a strategic intervention to disrupt existing travel patterns and behaviours, and encourage greater use of active transport overall^(89)^. Consideration of supportive services to limit reliance on volunteers, and structures to enhance implementation within a range of contexts may be beneficial for enhancing active school travel.

This review only considered interventions that aimed to promote the uptake of active travel. As a result, we did not consider interventions that aimed to reduce car use (often termed ‘stick’ interventions), that may have benefits for enabling modal shift to active travel. Recent reviews have suggested that the most effective interventions to decrease car use include congestion charging, parking and traffic controls, and limited traffic zones.^(90, 91)^ Congestion charging involves charging drivers to use the busiest roads or areas at the busiest times (often during peak periods), and has often been implemented in city centres. For example, congestion charging was implemented in Central London and resulted in a 33% reduction in motor vehicle traffic entering and leaving the congestion charging zone.^(92)^ Changes to parking and traffic controls includes the reduction in availability of on-street car parking and residential parking, parking pricing and workplace parking schemes. While evidence is limited, Knott and Sharp^(93)^ demonstrated that the introduction of free or subsidised workplace parking was associated with a higher proportion of motor vehicle trips. Limited traffic zones (including ‘low traffic neighbourhoods’ and ‘low emissions zones’) restrict traffic in specified areas, such as in city centres or residential areas), where non-residents and unauthorised vehicles are prohibited from driving at certain times. For example, low traffic neighbourhoods were implemented in London (primarily in residential areas using traffic management measures such as ‘modal filters’ to restrict general motor traffic and permit walking and cycling) and a 33% reduction in traffic volumes was observed.^(94)^ However, in the vast majority of cases, evaluations of interventions aimed to reduce car use have not reported on impacts to modal shift to walking and cycling.

Evidence also suggests that combinations (or ‘packages’) of ‘carrot’ and ‘stick’ interventions are more effective than individual interventions^(86)^. A systematic review by Xiao *et al.*^(20)^ identified that carrot and stick interventions or a mix of carrot and stick policy levers were more effective than carrot interventions alone at increasing active travel. An example of this is a ‘complete streets’ project that reallocates road space to ensure that people walking, bicycling, taking the bus and driving can more safely and easily get to where they need to go.^(95)^ Such a program includes carrot interventions, including protected bike lanes, wider and better lit sidewalks/footpaths, and landscaping, and stick interventions, such as a reduced number and/or width of motor vehicle traffic lanes, traffic calming and lower speed limits. In this systematic review, we were unable to account for combinations of interventions and future research should aim to understand which combinations of interventions are most effective at achieving modal shift to active travel. Additionally, we did not target interventions that had a sole aim to support the uptake of public transportation. Studies show that public transport users can get an additional 8 to 37 minutes of walking per day^(96)^ and therefore interventions to support modal shift from car travel to public transportation may confer substantial physical activity benefits.

### Unanswered questions and future research

This review highlighted need for substantial overhaul in how we conduct and measure the impact of interventions on modal shift to active transport. Poor consistency in reporting of outcomes, measurement of modal shift and study design limited interpretation of findings, and limits the usability of findings in further research. The active transport research field is multidisciplinary, cutting across multiple areas such as public health, transport engineering and urban planning. Robust methods for how randomised controlled trials and quasi-experimental studies have been developed within public health and medical research (used to limit bias) are needed to enhance the external and internal validity of findings. These methods are not common within other fields conducting evaluations of active transport interventions. Efforts to upskill other disciplines, such as transport engineering, in how to employ robust study designs are needed to ensure research efforts can be synthesised.

Studies included in this review aimed to increase the uptake of walking, cycling, or active transport overall, in an effort to shift people from sedentary modes such as the private motor vehicle. To understand which intervention types lead to people shifting from private motor vehicle use to sustainable and active modes such as walking and cycling, measures of modal shift are required. Measuring modal shift involves comparing the usage of different transport modes over time or in specific contexts.^(97)^ Most of the studies in this review did not measure modal shift; instead, they reported change in frequency, distance or duration of a single mode used. This is of concern as other measures do not accurately show if the observed change is due to a shift from a different mode, or an increase in the number of trips made by the measured mode. Modal shift can be determined in a variety of ways, including methods such as travel diaries, or GPS mode detection. A range of measures were used with the intent to determine modal shift within this review, including count data, pre-post surveys, GPS mode detection, travel diaries. Most studies measured the share of modes across total trips before and after the intervention occurred at an aggregate level (such as all trips an individual took across a two-week period), assuming changes in mode share were due to the intervention. Others measures were trip-specific and queried choice of one mode, such as the primary mode of travel to work. One study used both a stated-preference survey and a GPS smartphone app with mode detection, demonstrating that the stated-preference survey may have modestly overestimated modal shift to cycling compared to revealed preference methods like GPS-based mode detection^(98)^. While all modal shift measures included in the review provided some insight into intervention effectiveness, consideration of individual-level modal shift provided the most robust insight into how effective interventions were towards their aim of reducing private motor vehicle use. The field needs to rapidly move towards having a consistent set of outcomes measures when evaluating interventions to ensure the most robust measurement of effectiveness and to enable comparisons across studies.

Further to understanding modal shift effects, is the timing of measurement. Few studies in this review collected long-term longitudinal data, or conducted follow-for more than six months after intervention implementation. Travel behaviour is habitual, and difficult to change within a short time frame. Previous research that has used longitudinal data to explore the impacts of specific events on mode choice has demonstrated that modal shift often has a considerable time lag^(99)^. For example, Goodman et al. ^(100)^ demonstrated that changes in travel behaviour took up to two years following implementation of infrastructural interventions to support walking and cycling. Future studies should consider how data over longer time periods can be collected to capture the actual impact on travel behaviour resulting from the intervention, particularly in real-world environments. For example, hybrid effectiveness-implementation trials simultaneously evaluate effectiveness of the intervention on an outcome (for example, modal shift to active modes) and assess how the intervention is implemented within a real-world setting.^(101)^ This is particularly important in the context of active transport interventions where implementation often involves overcoming substantial and complex barriers.^(102)^

There was limited reporting of the impact of interventions on population sub-groups within this review. One study measured the impact of an intervention’s effectiveness by gender, demonstrating greater uptake by boys compared to girls within a school-setting.^(103)^ There are vast inequities in active travel participation, and particularly bike riding, where fewer women ride a bike in low-cycling countries, and most people riding are of high socio-economic status.^(104)^ Reporting of effectiveness that considers the impact the intervention has on particular population subgroups, particularly gender, is key to enabling understanding of how to overcome these inequities.

## Conclusion

This systematic review with meta-analysis of the effectiveness of interventions in increasing uptake in active transport provided critically needed evidence for which types of interventions warrant investment. Findings demonstrate that physical environmental restructure interventions, such as cycleways and traffic calming infrastructure, were most effective in increasing cycling frequency. Other intervention types, particularly persuasion, individually tailored behavioural programmes, and education, were also effective for increasing use of active modes. However, this body of research would benefit from more rigour in study design to limit lower quality evidence with the potential for bias. Further research to understand which combinations of intervention types, including disincentives to use other transport modes, are most effective for modal shift. Collectively, active transportation research needs to rapidly move towards more robust trials and evaluations with consistent outcome measures to improve the quality of evidence available and identify interventions (and combinations of interventions) that are most effective at increasing the uptake of active transportation, and in which populations and contexts.

## Data Availability

All data are available within supplementary materials and individual included studies.

## Notes

### Competing Interest Statement

The authors have declared no competing interest.

### Funding Statement

This study was funded by the British Columbia Centre for Disease Control.

### Author Declarations

This study only used human data that was available within published studies. All included studies are detailed within supplementary materials.

