## Supplementary material for "Effectiveness of interventions for modal shift to walking and bike riding: a systematic review with meta-analysis": Data Supplement 2

Data Supplement 2 – Search Strategies

OVID MEDLINE SEARCH STRATEGY

**Cycling/infrastructure specific:**

(((Environment Design/ OR Public Facilities/)  AND (Bicycling/)) OR ((bicycle* OR cycling  OR cyclist* OR biking OR bike ) *ADJ6* (lane* OR path* OR trail* OR infrastructure* OR planning OR design* OR network* OR space OR zone OR zones OR track* OR dedicate*) OR Bikeabilit* OR transport-infrastructure* OR (Cycle ADJ (lane OR path OR track OR lanes OR paths OR tracks OR network*)) OR (cycle ADJ3 (infrastructure* OR dedicate* OR path OR paths)) OR ((bicycle* OR cycling  OR cyclist* OR biking OR bike ) AND ((environment* OR infrastructur* OR neighbourhood*  OR neighborhood* OR roadway* OR boulevard* OR route*) ADJ6 (interven* OR change* OR plan* OR design* OR correlat* OR factor* OR determinant* OR difference* OR structure* OR characteristic* OR measure* OR built OR role OR condition* OR context* OR improve*)))).ab,ti.) **AND** (Leisure Activities/  OR recreation/ OR health promotion/ OR motor activity/ OR (((lifestyle OR life-style) *ADJ3* (modif* OR change* OR behav*  OR healthy)) OR ((physical* OR transport* OR travel*) *ADJ3* (activ* OR inactiv*)) OR  commut* OR leisur* OR recreat* OR ((transport* OR Prevalen* OR predictor* OR statistic* OR utilit* OR promot* OR increase* OR play OR "use" OR using) ADJ6 (cycling OR bicycling OR cyclist* OR biking OR bike)) OR (behav* ADJ3 (change* OR healthy)) OR (mode ADJ3 travel) OR (health ADJ3 promotion) OR (motor ADJ3 activit*)).ab,ti.) NOT (letter OR news OR comment OR editorial OR congresses OR abstracts).pt.

**Cycling:**

(Bicycling/ or Transportation/ or (cycling or bicycl* or riding or biking or commut* or bike or non-motoris* or non-motoriz* or e-bik* or (active adj2 travel) or (active adj2 transport) or (active adj2 commut*) or (ride adj3 school)).ti. **AND** (exp Health Promotion/ or Health Behavior/ or ((encourag* or promot* or intervention* or trial* or program* or policy* or campaign* or schem* or incentiv* or modif* or education* or behavio?r change) adj8 (cycling or bicycl* or riding or biking or commut* or bike or non-motori#* or e-bik* or (active adj5 (travel or transport or commut*)) or ((rid* or bik* or cycl*) adj5 (school or university or work or commut*)) or transport or travel*)).ti,ab,kf.) **AND** ((exp Controlled Clinical Trial/ or exp Epidemiologic Studies/ or (Comparative Study or Evaluation Study).pt. or (randomi#ed or randomly or time series or controlled before or non-randomi#ed or nonrandomi#ed or (before adj2 after) or (pre adj2 post) or control group* or ((case control or follow up or longitudinal or experimental) adj4 (study or studies or analys*))).ti,ab,kf. or (trial or study).ti.) not cross-sectional.ti,ab,kf.)

**Walking WITH TRANSPORT TERMS:**

(walking OR walk OR commut* OR pedestrian*  OR walk to school*) AND (encourag* OR promot* OR intervention* OR trial* OR program* OR policy* OR campaign* OR schem* OR incentiv* OR modif* OR education*) **AND** (encourag* or promot* or intervention* or trial* or program* or policy* or campaign* or schem* or incentiv* or modif* or education* or behaviour change or behavior change) adj8 ((walk adj3 commute) or (active adj2 travel) or (active adj2 transport) or (active adj2 commut*) or (walk adj3 work) or (walk adj3 school) or transport or travel*).ti,ab,kf.) **AND** ((exp Controlled Clinical Trial/ or exp Epidemiologic Studies/ or (Comparative Study or Evaluation Study).pt. or (randomi#ed or randomly or time series or controlled before or non-randomi#ed or nonrandomi#ed or (before adj2 after) or (pre adj2 post) or control group* or ((case control or follow up or longitudinal or experimental) adj4 (study or studies or analys*))).ti,ab,kf. or (trial or study).ti.) not cross-sectional.ti,ab,kf.)

**Walking WITHOUT TRANSPORT TERMS:**

(walking OR walk OR commut* OR pedestrian*  OR walk to school*) AND (encourag* OR promot* OR intervention* OR trial* OR program* OR policy* OR campaign* OR schem* OR incentiv* OR modif* OR education*) **AND** (encourag* or promot* or intervention* or trial* or program* or policy* or campaign* or schem* or incentiv* or modif* or education* or behaviour change or behavior change).ti,ab,kf.) **AND** ((exp Controlled Clinical Trial/ or exp Epidemiologic Studies/ or (Comparative Study or Evaluation Study).pt. or (randomi#ed or randomly or time series or controlled before or non-randomi#ed or nonrandomi#ed or (before adj2 after) or (pre adj2 post) or control group* or ((case control or follow up or longitudinal or experimental) adj4 (study or studies or analys*))).ti,ab,kf. or (trial or study).ti.) not cross-sectional.ti,ab,kf.)

WEB OF SCIENCE SEARCH STRATEGY

**Cycling:**

(cycling or bicycl* or riding or biking or commut* or bike or non-motoris* or non-motoriz* or e-bik* or (active NEAR/3 travel) or (active NEAR/3 transport) or (active NEAR/3 commut*) or (ride NEAR/3 school)) **AND** (encourag* or promot* or intervention* or trial* or program* or policy* or campaign* or schem* or incentiv* or modif* or education* or infrastructur* or “built environment*” or “behavior change” or “behaviour change”) NEAR/8 (“cycling” or bicycl* or “riding” or “biking” or commut* or “bike” or non-motoris* or non-motoriz* or e-bik* or transport* or travel*)

**Walking:**

(walking or walk or commut* or pedestrian* or (active NEAR/2 travel) or (active NEAR/2 transport) or (active NEAR/2 commut*) or (walk NEAR/3 school)) AND (encourag* or promot* or intervention* or trial* or program* or policy* or campaign* or schem* or incentiv* or modif* or education* or infrastructur* or “built environment*” or “behavior change” or “behaviour change”) NEAR/8 (walk* or transport* or travel*)

OVID PSYCHINFO SEARCH STRATEGY

**Cycling:**

(Cycling/ or Transportation/ or (cycling or bicycl* or riding or biking or commut* or bike or non-motoris* or non-motoriz* or e-bik* or (active adj2 travel) or (active adj2 transport) or (active adj2 commut*) or (ride adj3 school)).m_titl.) AND (exp Health Promotion/ or Health Behavior/ or Intervention/ or Urban environments/ or behavior change/ or Built Environment/ or ((encourag* or promot* or intervention* or trial* or program* or policy* or campaign* or schem* or incentiv* or modif* or education* or behavio?r change) adj8 (cycling or bicycl* or riding or biking or commut* or bike or non-motori#* or e-bik* or (active adj5 (travel or transport or commut*)) or ((rid* or bik* or cycl*) adj5 (school or university or work or commut*)) or transport or travel*)).m_titl.)

**Walking:**

Walking/ or Traveling/ or Transportation/ or (walking OR walk OR commut* OR pedestrian* OR walk to school*).m_titl.) **AND** (exp Health Promotion/ or Health Behavior/ or Intervention/ or Urban environments/ or behavior change/ or Built Environment/ or (encourag* or promot* or intervention* or trial* or program* or policy* or campaign* or schem* or incentiv* or modif* or education* or behaviour change or behavior change) adj8 ((walk adj3 commute) or (active adj2 travel) or (active adj2 transport) or (active adj2 commut*) or (walk adj3 work) or (walk adj3 school) or transport or travel*).m_titl.)
