## Supplementary material for "Effectiveness of interventions for modal shift to walking and bike riding: a systematic review with meta-analysis": Data Supplement 3

Data Supplement 3 – Study characteristics

Table 1. Study population descriptions

| **First author, Year of publication** | **Study population description** |
| --- | --- |
| Aittasalo, 2019 | Employees from small and middle-sized companies with more than 10 employees, located just outside the city centre of Tampere, Finland. |
| Aittasalo, 2012 | 20 office-based worksites with insufficiently physically active employees |
| Aldred, 2021 | Adults aged 16+ who lived in Outer London, both in mini-Holland and non-mini-Holland boroughs. |
| Aldred, 2019 | Adults aged 16+ who lived in Outer London, both in mini-Holland and non-mini-Holland boroughs. |
| Andersson, 2021 | Employees from a major company in Skövde |
| Aranda-Balboa, 2022 | Students from public secondary schools |
| Auchincloss, 2019 | Residents of the intervention and comparison areas |
| Audrey, 2019 | Employees from workplaces in seven urban areas in south-west of England and south Wales |
| Benton, 2021 | Adults observed using the intervention and comparison sites. |
| Bernstein, 2017 | Inactive adults in 2 lower-income neighbourhoods in Milwaukee, Wisconsin. One neighbourhood was largely a Latino community, while the other was primarily African American. |
| Biondi, 2022 | School children and their families residing in the intervention city Gdansk and control city Lodz |
| Bjornara, 2019 | Parents with children attending kindergarten and residing in Southern Norway. |
| Boarnet, 2005 | Parents of third- through fifth-grade children at ten schools that had a completed SR2S project nearby. |
| Brown, 2016 | Residents living Near (≤800 m) and Far (801–2000 m) from the complete street renovation in Salt Lake City, Utah |
| Brown, 2003 | University of California students and employees (faculty and staff) |
| Brownson, 2004 | Adults aged 18+ from communities that ranged in population from 2399 to 17,642 in Missouri “bootheel” region and in six comparison communities Arkansas and Tennessee. |
| Buckley, 2013 | Students from two elementary schools in Moscow, Idaho |
| Bungum, 2014 | Elementary school students in grades K-5 and located in Henderson, Nevada. |
| Cambra, 2020 | Adults in Lisbon, Portugal |
| Christiansen, 2014 | 5th-6th grade students from schools in the Region of Southern Denmark |
| Ciccone, 2021 | Adults (18+) from three major Norwegian cities (Oslo, Trondheim and Bergen). |
| Clark, 2014 | Trail users |
| Coffeng, 2014 | Office employees from a financial service provider |
| Coombes, 2016 | Primary school students in years 4 and 5 (aged 8-10yrs) |
| Crane, 2017 | Residents of the intervention and comparison areas |
| Crawford, 2013 | 5th and 6th grade primary school children |
| Dill, 2014 | Adults with children in Portland, OR |
| Ducheyne, 2014 | 4th grade students from primary schools in Flanders, Belgium |
| Duncan, 2011 | Grade 5-6 children from two diverse Auckland primary schools |
| Dunton, 2008 | Healthy, middle-aged women (ages 21–65 years) living in Northern and Southern California |
| Ek, 2020 | Adults from Stockholm County who were aged 20 to 65 years and had access to a smartphone. |
| Elinder, 2012 | School children aged 6–16 years in the middle-class municipality of Österåker (Stockholm, Sweden) |
| Felix, 2020 | Cyclists in Lisbon, Portugal |
| Fitzhugh, 2010 | Children, adolescents, and adults in free-living conditions within one experimental and two control neighbourhoods. |
| Folta, 2013 | Children enrolled in grades 1 to 3, typically ages 6 to 8 years, in the 10 Somerville public elementary schools |
| Frank, 2021 | Adults aged 18 and older who lived within 1 km of the greenway |
| Fyhri, 2015 | Members of the Norwegian Automobile Federation (NAF) living in the counties of Oslo and Akershus |
| Geng, 2020 | Car owners residing in Hefei city, China |
| Goodman, 2013 | Commuters living in the intervention and comparison tows |
| Goodman, 2016 | Children aged 10–11 years participating in the nationally-representative Millennium Cohort Study |
| Grimes, 2020 | College-aged students in an urban university in Kansas City, Missouri. |
| Grunseit, 2019 | Users of the intervention trail/residents living in close proximity to the new infrastructure |
| Gu, 2019 | Bike share users in Suzhou, China |
| Gutierrez, 2014 | School children and their parents |
| He, 2022 | Older adults (65+) residing in the intervention and control areas |
| Heelan, 2009 | Elementary school children from 3 schools in a small Midwestern community of 30,000 people in Nebraska. |
| Heesch, 2016 | Cyclists travelling in intervention/control area |
| Heinrich, 2011 | Students from the intervention and control schools |
| Hoelscher, 2016 | Fourth-grade children and their parents |
| Hong, 2016 | Residents of the intervention and comparison areas |
| Hosford, 2019 | Adults (18+) residing in cities that either had a newly implemented PBSP in 2013 (Chicago, New York), an existing PBSP (Boston, Montreal, Toronto), or no PBSP during the study period (Detroit, Philadelphia, Vancouver). |
| Hunter, 2021 | Adult residents (aged ≥16 years) who lived ≤1-mile radius of the greenway (intervention sample) and > 1-mile radius of the greenway (control sample) |
| Isensee, 2018 | 8th grade students from secondary schools in 6 districts of the federal state of Schleswig-Holstein, Germany |
| Jancey, 2011 | Older adults aged 65-74 years, residing in the Perth metropolitan area, Australia |
| Jones, 2012 | Residents of Beaconside (intervention neighbourhood) and Rickerscote (control neighbourhood) |
| Jordan, 2008 | First-, third-, and fifth-grade elementary school students and parents from four schools in Tooele County, UT. |
| Karpinski, 2021 | Users of the local bicycle sharing system 'BlueBikes' |
| Keall, 2022 | Residents aged 10yrs+ of the intervention and comparison areas |
| Keall, 2015 | People residing in New Plymouth, Hastings, Whanganui and Masterton |
| King, 2017 | Older adults at increased mobility disability risk |
| Kubota, 2020 | Older adults (aged 65-84 years) living in Oiso, Kanagawa Prefecture |
| Lambe, 2022 | 5th and 6th class students attending all primary schools in Waterford City. |
| Lambe, 2017 | 5th and 6th grade students in 3 Irish towns |
| Limb, 2020 | Adults (16+ years) seeking to move into social, intermediate, and market-rent East Village accommodation |
| Maca, 2020 | New users of the cycling smartphone application Cyclers, based in the Czech Republic |
| Mackey, 2019 | Community-dwelling men aged 60 years and older |
| Malakellis, 2017 | Secondary students aged 12–16 years, from government schools in Canberra ACT, Australia. |
| Mayer, 1982 | Residents of Foxridge Apartments. Individuals walking or bicycling in the direction of the Virginia Tech campus (Blacksburg, VA) on either of two bikeways during the observation periods |
| McDonald, 2014 | School-aged children in the District of Columbia (DC) and three states—Florida, Oregon, and Texas |
| McKee, 2007 | Two classes of primary 5 children and their families and teachers |
| McMinn, 2012 | Children from 5 elementary schools in Scotland. |
| Mendoza, 2017 | Fourth–fifth graders from four public schools serving low-income families in Seattle, WA |
| Mendoza, 2011 | 4th grade students from 8 low-income public elementary schools in Houston, Texas |
| Mendoza, 2009 | Ethnically diverse students in kindergarten-5th grade (aged 5–11 years) from urban, socioeconomically disadvantaged, public elementary schools in Seattle, Washington, USA |
| Merom, 2003 | Adults 18–55 years of age were randomly sampled from an “inner” area, residents within ≤1.5 km of the Trail, and an “outer” area, bike-owners only, 1.5–5 km from the Trail |
| Merom, 2007 | Inactive adults aged 30 – 65 years living in urban or rural regions of New South Wales (NSW) Australia |
| Mitra, 2021 | Adults (≥18 years old) living within 1 km of these 17 streets, who were identified based on their 6-digit postal code of residence. |
| Mutrie, 2002 | Employees from three city workplaces. |
| Olsen, 2016 | Residents located in: the local area surrounding the new motorway in the South of the city (intervention area), a residential area surrounding an existing motorway (the M8) in the East of the city (comparator area), and a residential area without a motorway in the North of the city (control area). |
| Ostergaard, 2015 | 4th and 5th grade students from public schools in three different regions in Denmark. |
| Petrunoff, 2016 | Clinical and non-clinical staff employed at Liverpool Hospital in Sydney, Australia. |
| Purath, 2005 | Sedentary female employees aged 18-69 years, who voluntarily attended a university-provided health screening as part of a wellness program. |
| Purath, 2004 | Sedentary female employees aged 18 to 65 who voluntarily attended a university-provided health screening as a part of a wellness program. |
| Rissel, 2010 | Residents from postcodes within two kilometres from the two bicycle paths who were >18 and spoke English. |
| Rissel, 2015 | Participants were aged 18–55 years, had ridden a bicycle in their life and had no current disability preventing them from riding. Resided either in intervention or comparison area. |
| Rothman, 2022 | Students from public elementary and secondary schools in the City of Toronto, Canada. |
| Rowland, 2003 | Primary school students in years 2 and 5 |
| Sahlqvist, 2019 | Primary school children in Victoria, Australia and their carers |
| Salinas, 2019 | Low-income, Mexican American women living in South Texas on the U.S.-Mexico border |
| Sersli, 2019 | Participants were eligible if they were aged 19 or older and had sufficient English (self-assessed) to complete the surveys (English-language only). |
| Simon, 2014 | 6th grade students from public middle schools of the Department of Bas-Rhin (Eastern France). |
| Simon, 2008 | 6th grade students from 8 public middle schools of the Department of Bas-Rhin (Eastern France). |
| Singh, 2009 | First-year students (aged 12-14 years) at prevocational secondary schools |
| Sirard, 2008 | Students from an elementary school in Menlo Park, California |
| Spears, 2016 | Residents from neighbourhoods within walking distance of new stations (the ‘experimental’ group), and comparison neighbourhoods with similar built environment and socio-demographic characteristics located further from the new stations (the ‘control’ group). |
| Stappers, 2021 | Participants (≥18 years) were part of one of three area-based exposure groups. The maximal exposure group lived in neighbourhoods directly bordering the Green Carpet. The minimal exposure group consisted of individuals living at the other side of the city, and the no exposure group consisted of individuals living in a nearby city. |
| Suchert, 2015 | 8th grade students from Germany |
| Sun, 2020 | Participants were 18–65 years of age, residents in the neighbourhood for at least three years and able to walk unassisted for at least 15 min |
| Sundfor, 2022 | People residing in Oslo, the capital of Norway. |
| Van Dyck, 2016 | Recently retired Belgian adults (>6 months, <5 years of retirement) |
| Van Stralen, 2010 | Older adults |
| Villa-Gonzalez, 2017 | Children aged 8 to 11 years from 3 schools in Salobreña, Huétor Vega and Santa Fe. |
| Villa-Gonzalez, 2016 | Children aged 8 to 11 years from 3 schools in Salobreña, Huétor Vega and Santa Fe. |
| Walsh, 2021 | Office workers aged ≥18 years working in the central business district of Brisbane, Australia. |
| Xiao, 2022 | Residents of Paris and Lyon |
| Xu, 2015 | 4th grade students from 8 urban primary schools in Nanjing City, China |

Table 2. Study characteristics

| **First Author, Year of Publication** | **Country** | **Mode of transport** | **Study Design** | **Sample Size** | **Intervention Type** | **Intervention Group Description** |
| --- | --- | --- | --- | --- | --- | --- |
| Aittasalo, 2019 ^49^ | Finland | AT | RCT | Phase 1: 1823 Phase 2: 826* | Education, Incentivisation, **Environmental restructuring (social),** Enablement | Employees from small and middle-sized companies with more than 10 employees, located just outside the city centre of Tampere, Finland. |
| Aittasalo, 2012 ^50^ | Finland | Walking | RCT | 241 | Education, Incentivisation, **Environmental restructuring (social),** Enablement | Volunteer and insufficiently physically active employees from 20 office-based worksites, recruited by 10 occupational health care units. |
| Aldred, 2021^51^ | England | AT | Controlled before-after study | 3435 | **Environmental restructuring (physical)** | Adults aged 16+ who lived in Outer London, both in mini-Holland and non-mini-Holland boroughs. |
| Aldred, 2019 ^52^ | England | AT | Controlled before-after study | 3435 | **Environmental restructuring (physical)** | Adults aged 16+ who lived in Outer London, both in mini-Holland and non-mini-Holland boroughs. |
| Andersson, 2021 ^53^ | Sweden | Cycling | RCT | 65 | **Enablement** | Employees from a major company in Skövde, Sweden. |
| Aranda-Balboa, 2022 ^54^ | Spain | Cycling & AT | RCT | 122 | Education, **Training** | Students from public secondary schools in Granada, Jaén and Valencia, Spain. |
| Auchincloss, 2019 ^55^ | USA | AT | Controlled before-after study | 175 | **Environmental restructuring (physical)** | Residents of the area local to the greenway construction; a poor, high-crime, predominantly African-American neighbourhood in Philadelphia, Pennsylvania. |
| Audrey, 2019 ^56^ | UK | AT | RCT | 654 | Education, **Persuasion,** Environmental restructuring (social) | Employees from workplaces in seven urban areas in south-west of England and south Wales. |
| Benton, 2021 ^57^ | England | Walking | Controlled before-after study | 116 | **Environmental restructuring (physical)** | Adults observed using the intervention sites in Greater Manchester, UK. |
| Bernstein, 2017 ^58^ | USA | Cycling | RCT | 38 | Persuasion, Training, **Enablement** | Inactive adults in 2 lower-income neighbourhoods in Milwaukee, Wisconsin. One neighbourhood was largely a Latino community, while the other was primarily African American. |
| Biondi, 2022 ^59^ | Poland | Cycling | Controlled before-after study | Not reported | **Persuasion** | School children and their families residing in the intervention city Gdansk. |
| Bjornara, 2019 ^60^ | Norway | Cycling | RCT | 36 | **Enablement** | Parents with children attending kindergarten and residing in Southern Norway. |
| Boarnet, 2005 ^61^ | USA | AT | Controlled before-after study | 862 | **Environmental restructuring (physical)** | Parents of third- through fifth-grade children at ten schools in California, USA that had a completed a Safe Routes to School project nearby. |
| Brown, 2016 ^62^ | USA | AT | Controlled before-after study | 910 | **Environmental restructuring (physical)** | Residents living Near (≤800 m) and Far (801–2000 m) from the complete street renovation in Salt Lake City, Utah |
| Brown, 2003 ^63^ | USA | AT | Controlled before-after study | 4565 | **Enablement** | University of California students and employees (faculty and staff). |
| Brownson, 2004 ^64^ | USA | Walking | Controlled before-after study | 1232 | **Environmental restructuring (social)** | Adults aged 18+ from communities that ranged in population from 2399 to 17,642 in Missouri Bootheel region. |
| Buckley, 2013 ^65^ | USA | AT | Controlled before-after study | Not reported | **Environmental restructuring (social)** | Students from two elementary schools in Moscow, Idaho. |
| Bungum, 2014 ^66^ | USA | AT | Controlled before-after study | 1336 | **Environmental restructuring (social)** | Elementary school students in grades K-5 and located in Henderson, Nevada. |
| Cambra, 2020 ^67^ | Portugal | Walking | Controlled before-after study | 802 | **Environmental restructuring (physical)** | Adults in Lisbon, Portugal. |
| Christiansen, 2014 ^68^ | Denmark | AT | RCT | 1279 | Environmental restructuring (physical), **Environmental restructuring (social)** | 5th-6th grade students from schools in the region of Southern Denmark. |
| Ciccone, 2021 ^69^ | Norway | Cycling | RCT | 486 | **Incentivisation** | Adults (18+) from three major Norwegian cities (Oslo, Trondheim and Bergen). |
| Clark, 2014 ^70^ | USA | AT | Controlled before-after study | Not reported | Education,  **Persuasion** | Users of six trails with signage additions in Southern Nevada. |
| Coffeng, 2014 ^71^ | Netherlands | AT | RCT | 412 | **Persuasion** | Office employees from a financial service provider who able to complete a Dutch (online) questionnaire and expect to be employed at the financial service provider during the complete follow-up period. |
| Coombes, 2016 ^72^ | England | AT | Controlled before-after study | 80 | **Incentivisation,**  Environmental restructuring (social) | Primary school students in years 4 and 5 (aged 8-10yrs) recruited via school in Norwich, UK. |
| Crane, 2017 ^73^ | Australia | Cycling | Controlled before-after study | 846 | **Environmental restructuring (physical)** | Residents of the intervention area; Sydney’s inner-city suburbs or Redfern and Waterloo. |
| Crawford, 2013 ^74^ | Australia | AT | Controlled before-after study | Not reported | **Environmental restructuring (physical),** Environmental restructuring (social) | 5th and 6th grade primary school children attending Government primary schools in metropolitan and regional areas of Victoria, Australia. |
| Dill, 2014 ^75^ | USA | AT | Controlled before-after study | 353 | **Environmental restructuring (physical)** | Adults with children in Portland, Oregon. |
| Ducheyne, 2014 ^76^ | Belgium | Cycling | RCT | 94 | **Training** | 4th grade students from primary schools in Flanders, Belgium. |
| Duncan, 2011 ^77^ | New Zealand | AT | RCT | 97 | **Education** | 5th and 6th grade children from two diverse Auckland primary schools. |
| Dunton, 2008 ^78^ | USA | Walking | RCT | 178 | **Persuasion** | Healthy, middle-aged women (aged 21–65 years) living in Northern and Southern California. |
| Ek, 2020 ^79^ | Sweden | AT | RCT | 252 | **Persuasion**,  Incentivisation | Adults from Stockholm County who were aged 20 to 65 years and had access to a smartphone. |
| Elinder, 2012 ^80^ | Sweden | AT | Controlled before-after study | 813 | Persuasion,  **Environmental restructuring (social)** | School children aged 6–16 years in the middle-class municipality of Österåker (Stockholm, Sweden). |
| Felix, 2020 ^81^ | Portugal | Cycling | Controlled before-after study | 6,414 | **Environmental restructuring (physical),**  Enablement | Cyclists in Lisbon, Portugal. |
| Fitzhugh, 2010 ^82^ | USA | AT | Controlled before-after study | Not reported | **Environmental restructuring (physical)** | Children, adolescents, and adults in free-living conditions in a defined intervention neighbourhood in Knoxville–Knox County, Tennessee. |
| Folta, 2013 ^83^ | USA | Walking | Controlled before-after study | 454 | Education,  Incentivisation,  **Environmental restructuring (social)** | Children enrolled in grades 1 to 3, typically aged 6 to 8 years, in the 10 Somerville public elementary schools in Massachusetts, USA. |
| Frank, 2021 ^84^ | Canada | Cycling | Controlled before-after study | 524 | **Environmental restructuring (physical)** | Adults aged 18 and older who lived within 1 km of the greenway in Vancouver, Canada. |
| Fyhri, 2015 ^85^ | Norway | Cycling | RCT | 226 | **Enablement** | Members of the Norwegian Automobile Federation (NAF) living in the counties of Oslo and Akershus. |
| Geng, 2020 ^48^ | China | AT | Controlled before-after study | 146 | **Education,**  Incentivisation | Car owners residing in Hefei city, China. |
| Goodman, 2013 ^86^ | England | Cycling | Controlled before-after study | Not reported | Training,  **Environmental restructuring (physical),** Environmental restructuring (social) | Commuters living in one of the 18 intervention towns (Darlington, Derby, Brighton Gove, Aylesbusy. Exeter, Lancaster Morcambe, York, Cambridge (and surrounding areas), Colchester, Southend-On-Sea, Leighton Buzzards, Woking/Byfleet/Chobham, Bristol (sections), Shrewsbury, Stoke-on-Trent (sections), Chester, Southport, Blackpool). |
| Goodman, 2016 ^87^ | England | Cycling | Controlled before-after study | 3336 | **Training** | Children aged 10–11 years participating in the nationally-representative Millennium Cohort Study. |
| Grimes, 2020 ^88^ | USA | Cycling | RCT | 53 | **Enablement** | College-aged students in an urban university in Kansas City, Missouri. |
| Grunseit, 2019 ^89^ | Australia | AT | ITS | 249 | **Environmental restructuring (physical)** | Users of the intervention trail and residents living in close proximity to the new infrastructure in Northern Sydney. |
| Gu, 2019 ^90^ | China | Cycling | Controlled before-after study | 275 | **Environmental restructuring (physical)** | Bike share users in Suzhou, China. |
| Gutierrez, 2014 ^91^ | USA | AT | Controlled before-after study | Not reported | Environmental restructuring (physical), **Environmental restructuring (social)** | School children and their parents in Toronto, Canada. |
| He, 2022 ^92^ | China | Walking | Controlled before-after study | 449 | **Environmental restructuring (physical)** | Older adults (65+) residing in the intervention areas of Hong Kong. |
| Heelan, 2009 ^93^ | USA | Walking | Controlled before-after study | 324 | **Environmental restructuring (social)** | Elementary school children from 3 schools in a small Midwestern community of 30,000 people in Nebraska. |
| Heesch, 2016 ^94^ | Australia | Cycling | Controlled before-after study | 301 | **Environmental restructuring (physical)** | Cyclists travelling in intervention areas of Brisbane, Australia. |
| Heinrich, 2011 ^95^ | USA | AT | Controlled before-after study | 1648 | **Environmental restructuring (physical),** Environmental restructuring (social) | Students attending one of the intervention schools in Hawai‘i. Schools were eligible for inclusion if (1) they have ≥35% of students who qualified for free and reduced school lunch; (2) they were willing to fulfil study requirements; and (3) they were rural (<20% of student lived within 1-mile) or neighborhood (>60% of students lived within 1-mile). |
| Hoelscher, 2016 ^96^ | USA | AT | Controlled before-after study | 78 schools | **Environmental restructuring (physical),** Environmental restructuring (social) | Fourth-grade children and their parents attending randomly selected primary schools in Texas. |
| Hong, 2016 ^97^ | USA | Walking/AT | Controlled before-after study | 204 | **Environmental restructuring (physical)** | Residents of study neighbourhood, located along the Expo light rail line in south Los Angeles, California. |
| Hosford, 2019 ^98^ | USA & Canada | Cycling | Controlled before-after study | Not reported | **Enablement** | Adults (18+) residing in cities that either had a newly implemented public bike share (PBSP) in 2013 (Chicago, New York), an existing PBSP (Boston, Montreal, Toronto). |
| Hunter, 2021 ^99^ | Northern Ireland | AT | Controlled before-after study | 1205 | **Environmental restructuring (physical)** | Adult residents (aged ≥16 years) who lived ≤1-mile radius of the greenway. |
| Isensee, 2018 ^100^ | Germany | AT | RCT | 1020 | **Incentivisation** | 8th grade students from secondary schools in 6 districts of the federal state of Schleswig-Holstein, Germany. |
| Jancey, 2011 ^101^ | Australia | Walking | Controlled before-after study | 573 | Persuasion,  **Environmental restructuring (social)** | Older adults aged 65-74 years, residing in the Perth metropolitan area, Australia. |
| Jones, 2012 ^102^ | UK | Cycling | Controlled before-after study | 205 | **Environmental restructuring (physical)** | Residents of the intervention neighbourhood, Beaconside UK. |
| Jordan, 2008 ^103^ | USA | AT | Controlled before-after study | 578 | **Environmental restructuring (social)** | First-, third-, and fifth-grade elementary school students and parents from four schools in Tooele County, Utah. |
| Karpinski, 2021 ^104^ | USA | Cycling | Controlled before-after study | Not reported | **Environmental restructuring (physical)** | Users of the local bicycle sharing system 'BlueBikes' in Boston, Massachusetts. |
| Keall, 2022 ^105^ | New Zealand | AT | Controlled before-after study | 2,500 | Persuasion,  **Environmental restructuring (physical)** | People aged 10+ residing in New Plymouth and Hasting, New Zealand. |
| Keall, 2015 ^106^ | New Zealand | AT | Controlled before-after study | 749 | **Environmental restructuring (physical),** Environmental restructuring (social) | People residing in New Plymouth, Hastings, Whanganui and Masterton, New Zealand. |
| King, 2017 ^107^ | USA | Walking | RCT | 400 | **Persuasion** | Older adults at increased mobility disability risk; ages 70-89 years; from Dallas, Texas, San Francisco California, Pittsburgh, Pennsylvania, and Winston-Salem, North Carolina). |
| Kubota, 2020 ^108^ | Japan | Walking | Controlled before-after study | 650 | Education,  **Environmental restructuring (social)** | Older adults (aged 65-84 years) living in Oiso, Kanagawa Prefecture, Japan. |
| Lambe, 2022 ^109^ | Ireland | AT | Controlled before-after study | 1289 | Incentivisation,  **Environmental restructuring (social)** | 5th and 6th class students attending all primary schools in Waterford City, Ireland. |
| Lambe, 2017 ^110^ | Ireland | AT | Controlled before-after study | 1457 | Education,  **Environmental restructuring (physical),** Environmental restructuring (social) | 5th and 6th grade students in two intervention towns in Ireland. |
| Limb, 2020 ^111^ | England | AT | Controlled before after study | 578 | **Environmental restructuring (physical)** | Adults (16+ years) seeking to move into social, intermediate, and market-rent East Village accommodation in London. |
| Maca, 2020 ^112^ | Czech Republic | Cycling | RCT | 482 | **Incentivisation** | New users of the cycling smartphone application Cyclers, based in the Czech Republic. |
| Mackey, 2019 ^113^ | Canada | AT | RCT | 58 | **Persuasion**,  Enablement | Community-dwelling men aged 60 years and older in Metro Vancouver who wanted to be more physically active, had not participated in any strength training or aerobic exercise classes in the past 3 months, and did not have plans to be out of town for ≥7 days during the study. |
| Malakellis, 2017 ^114^ | Australia | Walking | Controlled before-after study | 656 | Environmental restructuring (physical), **Environmental restructuring (social),** Enablement | Secondary students aged 12–16 years, from government schools in Canberra Australian Capital Territory, Australia. |
| Mayer, 1982 ^115^ | USA | AT | Controlled before-after study | 464 | **Incentivisation** | Residents of Foxridge Apartments. Individuals walking or bicycling in the direction of the Virginia Tech campus (Blacksburg, VA) on either of two bikeways during the observation periods. |
| McDonald, 2014 ^116^ | USA | AT | Controlled before-after study | 26,231 | **Environmental restructuring (physical),** Environmental restructuring (social) | School-aged children in the District of Columbia and three states: Florida, Oregon, and Texas. |
| McKee, 2007 ^117^ | Scotland | Walking | Controlled before-after study | 60 | **Environmental restructuring (social)** | Children who lived within the statutory walking distance ( West Dunbartonshire, Scotland) of the study primary school in West Dunbartonshire, Scotland. |
| McMinn, 2012 ^118^ | Scotland | AT | Controlled before-after study | 166 | **Environmental restructuring (social)** | Children from 5 elementary schools in Scotland. |
| Mendoza, 2017 ^119^ | USA | Cycling | RCT | 54 | **Environmental restructuring (social),** Enablement | Fourth–fifth graders from four public schools serving low-income families in Seattle, WA. |
| Mendoza, 2011 ^120^ | USA | AT | RCT | 149 | **Environmental restructuring (social)** | 4th grade students from 8 low-income public elementary schools in Houston, Texas. |
| Mendoza, 2009 ^121^ | USA | Walking | Controlled before-after study | 820 | **Environmental restructuring (social)** | Ethnically diverse students in kindergarten-5th grade (aged 5–11 years) from urban, socioeconomically disadvantaged, public elementary schools in Seattle, Washington, USA. |
| Merom, 2003 ^122^ | Australia | AT | Controlled before-after study | 568 | **Persuasion**,  Environmental restructuring (physical) | Adults 18–55 years of age were randomly sampled from an “inner” area, residents within ≤1.5 km of the Trail, and an “outer” area, bike-owners only, 1.5–5 km from the Trail. |
| Merom, 2007 ^123^ | Australia | Walking | RCT | 369 | **Persuasion** | Inactive adults aged 30 – 65 years living in urban or rural regions of New South Wales, Australia. |
| Mitra, 2021 ^124^ | Canada | Cycling | Controlled before-after study | 1640 | **Environmental restructuring (physical)** | Adults (≥18 years old) living within 1 km of these 17 streets, who were identified based on their 6-digit postal code of residence. |
| Mutrie, 2002 ^125^ | Scotland, UK | AT & Walking | RCT | 295 | Education,  **Persuasion,**  Enablement | Employees from three city workplaces in Glasgow, Scotland. |
| Olsen, 2016 ^126^ | Scotland, UK | AT | Controlled before-after study | 1453 | **Environmental restructuring (physical)** | Residents located in: the local area surrounding the new motorway in the south of Glasgow, Scotland. |
| Ostergaard, 2015 ^127^ | Denmark | Cycling | Controlled before-after study | 2401 | Incentivisation,  Training,  **Environmental restructuring (physical**) | 4th and 5th grade students from public schools in three different regions in Denmark. |
| Petrunoff, 2016 ^128^ | Australia | AT | ITS | 804 | Incentivisation,  Environmental restructuring (physical), **Environmental restructuring (social)** | Clinical and non-clinical staff employed at Liverpool Hospital in Sydney, Australia. |
| Purath, 2005 ^129^ | USA | Walking | RCT | 287 | **Persuasion** | Sedentary female employees aged 18-69 years, who voluntarily attended a university-provided health screening as part of a wellness program. |
| Purath, 2004 ^130^ | USA | Walking | RCT | 271 | **Persuasion** | Sedentary female employees aged 18 to 65 who voluntarily attended a university-provided health screening as a part of a wellness program. |
| Rissel, 2010 ^131^ | Australia | Cycling | Controlled before-after study | 1450 | Education,  Persuasion,  Training,  **Environmental restructuring (social)** | Residents from postcodes within two kilometres from the two bicycle paths who were >18 and spoke English. |
| Rissel, 2015 ^132^ | Australia | AT | Controlled before-after study | 846 | **Environmental restructuring (physical)** | Participants were aged 18–55 years, had ridden a bicycle in their life and had no current disability preventing them from riding residing in the intervention area in inner-Sydney. |
| Rothman, 2022 ^133^ | Canada | AT | Controlled before-after study | 885 | **Environmental restructuring (physical)** | Students from public elementary and secondary schools in Toronto, Canada. |
| Rowland, 2003 ^134^ | England | AT | RCT | 1386 | Persuasion,  **Environmental restructuring (social)** | Primary school students in years 2 and 5 attending intervention schools in the London boroughs of Camden and Islington. |
| Sahlqvist, 2019 ^135^ | Australia | AT | Controlled before-after study | 813 | Education,  Incentivisation,  **Environmental restructuring (social)** | Primary school children in Victoria, Australia and their carer’s. |
| Salinas, 2019 ^136^ | USA & Mexico | Walking | RCT | 620 | **Education**,  Environmental restructuring (social) | Low-income, Mexican American women living in South Texas on the U.S.-Mexico border. |
| Sersli, 2019 ^137^ | Canada | Cycling | Controlled before-after study | 178 | **Training** | Participants were eligible if they were aged 19 or older and had sufficient English (self-assessed) to complete the surveys (English-language only). |
| Simon, 2014 ^138^ | France | AT | RCT | 1048 | Education,  **Environmental restructuring (social)** | 6th grade students from public middle schools of the Department of Bas-Rhin (Eastern France). |
| Simon, 2008 ^139^ | France | AT | RCT | 954 | Education,  **Environmental restructuring (social)** | 6th grade students from 8 public middle schools of the Department of Bas-Rhin (Eastern France). |
| Singh, 2009 ^140^ | Netherlands | AT | RCT | 1108 | **Education**,  Environmental restructuring (social) | First-year students (aged 12-14 years) at prevocational secondary schools. |
| Sirard, 2008 ^141^ | USA | Walking | RCT | 11 | **Environmental restructuring (social)** | Students from an elementary school in Menlo Park, California. |
| Spears, 2016 ^142^ | USA | AT | Controlled before-after study | 285 | **Environmental restructuring (physical)** | Residents from neighbourhoods within walking distance of new stations (the ‘experimental’ group), and comparison neighbourhoods with similar built environment and socio-demographic characteristics located further from the new stations (the ‘control’ group). |
| Stappers, 2021 ^143^ | Netherlands | AT | Controlled before-after study | 642 | **Environmental restructuring (physical)** | Participants (≥18 years) were part of one of three area-based exposure groups. The maximal exposure group lived in neighbourhoods directly bordering the Green Carpet. The minimal exposure group consisted of individuals living at the other side of the city, and the no exposure group consisted of individuals living in a nearby city. |
| Suchert, 2015 ^144^ | Germany | AT | RCT | 1162 | **Incentivisation** | 8th grade students from Germany. |
| Sun, 2020 ^145^ | China | AT | Controlled before-after study | 5436 | **Environmental restructuring (physical)** | Participants were 18–65 years of age, residents in the neighbourhood for at least three years and able to walk unassisted for at least 15 min. |
| Sundfor, 2022 ^146^ | Norway | AT | Controlled before-after study | 1261 | **Incentivisation** | People residing in Oslo, Norway. |
| Van Dyck, 2016 ^147^ | Belgium | Walking | RCT | 284 | Education,  **Persuasion** | Recently retired Belgian adults (>6 months, <5 years of retirement). |
| van Stralen, 2010 ^148^ | Netherlands | AT | RCT | 1971 | Education, **Persuasion** | Adults aged ≥50, living in the Netherlands. |
| Villa-Gonzalez, 2017 ^149^ | Spain | AT | Controlled before-after study | 469 | **Education** | Children aged 8 to 11 years from 3 schools in Salobreña, Huétor Vega and Santa Fe, Spain. |
| Villa-Gonzalez, 2016 ^150^ | Spain | AT | Controlled before-after study | 206 | **Education** | Children aged 8 to 11 years from 3 schools in Salobreña, Huétor Vega and Santa Fe, Spain. |
| Walsh, 2021 ^151^ | Australia | AT | Controlled before-after study | 68 | **Persuasion,**  Education, Modelling | Office workers aged ≥18 years working in the central business district of Brisbane, Australia. |
| Xiao, 2022 ^152^ | France | Cycling | Controlled before-after study | Not reported | **Environmental restructuring (physical)** | Residents of Paris and Lyon, France. |
| Xu, 2015 ^153^ | China | AT | RCT | 1182 | Education,  **Environmental restructuring (social)** | 4th grade students from 8 urban primary schools in Nanjing City, China. |

Table 3. Intervention descriptions

| **First author, Year of publication** | **Intervention type (Kelly)** | **Intervention description** | **Intervention summary** | **Primary outcomes** |
| --- | --- | --- | --- | --- |
| Geng, 2020^48^ | **Education,**  Incentivisation | Periodic educational messages in a WeChat group containing: 1) Environmental information about the carbon emissions from a fossil fuel car at different speeds, a comparison to buses and rail transit and information about the contribution of car exhaust to air pollution.  2) Health information informing participants that the World Health Organisation reports reductions in health conditions such as cancers, CVD, dementia, obesity and mental health conditions that can be obtained by walking or bike riding for 30 minutes per day. 3) The same information as message two except specifying 45 minutes per day instead of 30. | Text message educational program | Walking duration Walking frequency Cycling duration Cycling frequency Modal shift duration Modal shift frequency |
| Salinas, 2019^136^ | **Education**,  Environmental restructuring (social) | 16-week long intervention that included educational classes and promotor-led PA sessions (i.e. aerobic or walking groups). | Group educational program | Walking duration Cycling duration |
| Duncan, 2011^77^ | **Education** | Six-week ‘Healthy Homework’ programme and complementary teaching resource to improve physical activity and dietary behaviours in children. | School based educational program | Step count, AT frequency |
| Singh, 2009^140^ | **Education**,  Environmental restructuring (social) | An interdisciplinary program with an adapted curriculum for 11 lessons in biology and physical education and environmental change options. | School based educational program | AT duration |
| Villa-Gonzalez, 2016^150^ | **Education** | Monthly 60-120-minute activities during school hours in addition to regular Physical Education lessons. Included questionnaire, story reading, knowledge on environmental characteristics, road safety, behaviours on the street, traditional games. Lessons targeted individual factors such as safety perceptions of walking to school and attitudes towards independence and motivations to walk. | School based educational program | AT frequency Modal shift frequency |
| Villa-Gonzalez, 2017^149^ | **Education** | Monthly 60-120-minute activities during school hours in addition to regular Physical Education lessons. Included questionnaire, story reading, knowledge on environmental characteristics, road safety, behaviours on the street, traditional games. Lessons targeted individual factors such as safety perceptions of walking to school and attitudes towards independence and motivations to walk. | School based educational program | AT frequency Modal shift frequency |
| Clark, 2014^70^ | Education,  **Persuasion** | Marketing campaign promoting trail use. Way-finding and incremental distance signage added to selected trails. | Campaign with signage and wayfinding | Pedestrian counts |
| Audrey, 2019^56^ | Education, **Persuasion,** Environmental restructuring (social) | Walk to Work Event: providing information about the benefits of walking to work; encouraging intention formation; identifying barriers and solutions; goal setting; self-monitoring (with travel diaries and optional pedometers); providing general encouragement; identifying social support; reviewing goals, and; relapse prevention. | Individually tailored behavioural programme | MVPA |
| Biondi, 2022^59^ | **Persuasion** | Cycling May: a public campaign aimed at increasing the usage of bicycles, primarily targeting children. | Individually tailored behavioural programme | Bicycle counts |
| Coffeng, 2014^71^ | **Persuasion** | Group motivational interviewing/counselling for employees on a worksite. | Individually tailored behavioural programme | MVPA |
| Dunton, 2008^78^ | **Persuasion** | Access to a tailored website and weekly emails containing links to a webpage with an interactive information tailoring tool to promote physical activity. | Individually tailored behavioural programme | Walking duration  MVPA |
| Ek, 2020^79^ | **Persuasion**,  Incentivisation | TravelVu Plus: 3-month behaviour change program aimed at increasing physical activity through active transport delivered as extra features to the TravelVU app. | Individually tailored behavioural programme | MVPA Walking duration Cycling duration |
| Mackey, 2019^113^ | **Persuasion**,  Enablement | Trained activity coaches delivered: (a) one-on-one participant consultations to develop personal action plans for physical activity and active transportation, (b) monthly group-based motivational meetings, (c) weekly telephone support, (d) complimentary recreation and transit passes, and (e) pedometers and diaries for self-monitoring. | Individually tailored behavioural programme | Walking frequency AT frequency Modal shift frequency |
| Merom, 2007^123^ | **Persuasion** | Intervention 1: theoretically based self-help walking program and weekly diaries (sent by mail).   Intervention 2: the same walking program with a pedometer also by mail. | Individually tailored behavioural programme | Walking duration Walking frequency |
| Mutrie, 2002^125^ | Education,  **Persuasion,**  Enablement | Booklet with written interactive materials based on the transtheoretical model of behaviour change, educational, and practical information on: choosing routes, maintaining personal safety, shower and safe cycle storage information, and useful contacts. The pack also included an activity diary in the form of a wall chart, a workplace map, distances from local stations, local cycle retailers and outdoor shops, contacts for relevant organisations, local maps, and reflective safety accessories. | Individually tailored behavioural programme | Walking duration |
| Purath, 2004^130^ | **Persuasion** | Brief counselling intervention, tailored to Stage of Change and designed to increase physical activity. | Individually tailored behavioural programme | MVPA  Walking duration |
| Purath, 2005^129^ | **Persuasion** | Brief counselling tailored to participant's State of Change, focused on women. | Individually tailored behavioural programme | MVPA  Walking duration |
| Van Dyck, 2016^147^ | Education,  **Persuasion** | Self-regulation eHealth intervention ‘MyPlan1.0.’ which provided personalised feedback on how to meet health guidelines of 150 minutes per week of physical activity. | Individually tailored behavioural programme | Walking duration Cycling duration |
| Van Stralen, 2010^148^ | Education,  **Persuasion** | Participants received three tailored letters including personalized physical activity advice.  Environmentally tailored intervention: participants received the same tailored information as the basic tailored intervention participants but additionally received tailored information about physical activity opportunities in their specific environment. | Individually tailored behavioural programme | Walking duration Cycling duration |
| Walsh, 2021^151^ | **Persuasion,**  Education,  Modelling | Commuter Choices: a six-week program including four lunchtime seminars, goal-setting, personalised journey plans, a buddy system and online information. | Individually tailored behavioural programme | AT frequency |
| King, 2017^107^ | **Persuasion** | A combination of centre-based (supervised) and home-based physical activity, with a goal of 150 min of walking/week to target aerobic activities. | Individually tailored behavioural programme with supervised activity | Walking duration |
| Ciccone, 2021^69^ | **Incentivisation** | 1. Mobile app where participants were paid 2 NOK for each kilometre cycled and registered on the app.   2. Mobile app where participants would gain one lottery ticket for each kilometre cycled and registered on the app. At the end of the experiment one lottery ticket would be drawn and one winner would receive NOK 9000.  3. The same as treatment 2 with the addition that the lottery winner must have registered at least one cycling trip on a randomly selected day to receive the money. If they had not logged a cycling trip on the randomly selected day, they would receive an email that they had won the lottery but would not receive the money as they hadn’t logged a trip on the randomly selected day. This additional risk of losing the money was designed to evoke regret aversion. | App based financial rewards system for participation | Cycling frequency Cycling distance |
| Maca, 2020^112^ | **Incentivisation** | The 'Cyclers' smartphone app consists of a built-in system of points, badges, leader-boards, and challenges, combined with personalized push and in-app notifications. Financial rewards were offered to participants. | App based financial rewards system for participation | Cycling frequency Cycling distance |
| Mayer, 1982^115^ | **Incentivisation** | Contest entry for travelling on bike path connecting Virginia Tech campus to Foxridge Apartment complex. | Lottery contest for participation | Cycling frequency Modal shift frequency |
| Isensee, 2018^100^ | **Incentivisation** | School-based physical activity program that integrated different behaviour change strategies such as self-monitoring, goal setting, class competitions, and social support with pedometer use. | School based competition for participation | MVPA Walking duration |
| Suchert, 2015^144^ | **Incentivisation** | Students received pedometers and took part in a class competition over a time period of 12 weeks. Classes with the most steps and best creative ideas to promote physical activity in everyday life were awarded. | School based competition for participation | AT duration |
| Sundfor, 2022^154^ | **Incentivisation** | Subsidy program for e-bike purchasers whereby the first 1000 applicants were awarded a subsidy of 25 per cent of the cost up to a maximum of 5000 NOK. | Financial subsidy for e-bike purchase | Walking frequency Cycling frequency Modal shift frequency |
| Aranda-Balboa, 2022 ^54^ | Education, **Training** | School-based intervention to promote cycling to school within Physical Education sessions, including an education session and two bicycling training sessions. | School based cycle training | Cycling frequency AT frequency |
| Ducheyne, 2014^76^ | **Training** | Cycle training course (four sessions) and parental involvement in homework tasks. Homework tasks included annotating a picture of a bicycle to indicate the mandatory requirements, indicating the most dangerous traffic spots near the school on a map, verifying their bicycle was in line with legal requirements and indicating the meanings of different road signs. Homework tasks specified they should be completed with the child’s parents. | School based cycle training | Cycling frequency |
| Goodman, 2016^87^ | **Training** | Bikeability cycle training scheme: aims to give children practical skills and understanding about how to cycle on today’s roads. | Cycle training scheme for children | Cycling frequency |
| Sersli, 2019^137^ | **Training** | Bicycle courses delivered by accredited instructors, 2 to 4.5 hours in duration, aimed to increase participant comfort level to ride on residential and urban streets through teaching in-person and on-road traffic handling skills. | Bicycle skills training | Cycling frequency |
| Aldred, 2021 & 2019^51, 52^ | **Environmental restructuring (physical)** | Mini Hollands programme: major investments in active travel infrastructure in three Outer London boroughs. | Supportive active transport infrastructure (e.g. cycleways and traffic calming) | Walking frequency Cycling frequency AT frequency |
| Auchincloss, 2019^55^ | **Environmental restructuring (physical)** | 1.5-mile urban greenway constructed along arterial streets in a neighbourhood in Philadelphia, Pennsylvania. | Supportive active transport infrastructure (e.g. cycleways and traffic calming) | Cycling frequency AT frequency |
| Benton, 2021^57^ | **Environmental restructuring (physical)** | Green space improvements along an urban canal. | Supportive active transport infrastructure (e.g. cycleways and traffic calming) | Physical activity |
| Boarnet, 2005^61^ | **Environmental restructuring (physical)** | Safe Routes to School (SR2S) California program: pedestrian and bicycle infrastructure improvements including installation or widening of bicycle lanes and crosswalks, installation of sidewalks and kerb ramps. | Supportive active transport infrastructure (e.g. cycleways and traffic calming) | AT frequency |
| Brown, 2016^62^ | **Environmental restructuring (physical)** | Complete street intervention including new light rail, bike lanes, and better sidewalks in Salt Lake City, Utah. | Supportive active transport infrastructure (e.g. cycleways and traffic calming). and New public transport service (e.g. new train line or light rail service) | Walking frequency Cycling frequency Modal shift frequency |
| Cambra, 2020^67^ | **Environmental restructuring (physical)** | The Eixo Central project aimed at improving walking conditions by changing physical factors (more green space, wider pedestrian footpaths, fewer lanes for motorised transport) in three sites. | Supportive active transport infrastructure (e.g. cycleways and traffic calming) | Pedestrian counts |
| Crane, 2017^73^ | **Environmental restructuring (physical)** | A 2.4km bi-directional protected cycleway was built through two inner city Sydney suburbs. | Supportive active transport infrastructure (e.g. cycleways and traffic calming) | Cycling frequency Cycling duration Bicycle counts Modal shift |
| Crawford, 2013^74^ | **Environmental restructuring (physical),** Environmental restructuring (social) | Ride2School program: Ride2School Day, mapping of safe routes to school, infrastructure improvements (e.g., bicycle storage funding and raised pedestrian crossings). | Supportive active transport infrastructure (e.g. cycleways and traffic calming) | AT frequency |
| Dill, 2014^75^ | **Environmental restructuring (physical)** | Bicycle boulevard installation in Portland, USA. | Supportive active transport infrastructure (e.g. cycleways and traffic calming) | AT frequency |
| Felix, 2020^81^ | **Environmental restructuring (physical),**  Enablement | Development of 100 km of dedicated cycling infrastructure, addition of a bike-sharing system of 1,400 bicycles, with 70% of the fleet as e-bikes. | Supportive active transport infrastructure (e.g. cycleways and traffic calming) | Bicycle counts |
| Fitzhugh, 2010^82^ | **Environmental restructuring (physical)** | Urban greenway/trail and improved pedestrian infrastructure. | Supportive active transport infrastructure (e.g. cycleways and traffic calming) | AT frequency |
| Frank, 2021^84^ | **Environmental restructuring (physical)** | Construction of an urban greenway. | Supportive active transport infrastructure (e.g. cycleways and traffic calming) | Cycling frequency Modal shift frequency |
| Goodman, 2013^86^ | Training,  **Environmental restructuring (physical),** Environmental restructuring (social) | Town-wide cycling initiatives in six Cycling Demonstration Towns and 12 Cycling Cities and Towns. The initiatives involved capital investment (e.g. cycle lanes) and revenue investment (e.g. cycle training), tailored to each town. | Supportive active transport infrastructure (e.g. cycleways and traffic calming) | Cycling frequency |
| Grunseit, 2019^89^ | **Environmental restructuring (physical)** | Development of a recreational walking and cycling loop trail. | Supportive active transport infrastructure (e.g. cycleways and traffic calming) | Pedestrian counts Bicycle counts |
| Heesch, 2016^94^ | **Environmental restructuring (physical)** | A new segment of a dedicated bikeway that links southern suburbs with Brisbane city centre, Australia. | Supportive active transport infrastructure (e.g. cycleways and traffic calming) | Cycling distance  Bicycle counts |
| Heinrich, 2011^95^ | **Environmental restructuring (physical),** Environmental restructuring (social) | State-wide policy, locally-based Safe Routes to School (SRTS) programs and bicycle and pedestrian planning initiatives. | Supportive active transport infrastructure (e.g. cycleways and traffic calming) | Walking frequency Cycling frequency Modal shift |
| Hoelscher, 2016^96^ | **Environmental restructuring (physical),** Environmental restructuring (social) | Safe Routes to School: Infrastructure schools applied for funding for building projects or improvements, non-infrastructure schools applied for funding for developing or implementing a plan initiative | Supportive active transport infrastructure (e.g. cycleways and traffic calming) | AT frequency |
| Hunter, 2021^99^ | **Environmental restructuring (physical)** | Provision of a 9 km urban greenway along the course of 3 rivers; 16 km of new or improved foot and cycle paths; development of a new civic square; development of 8 tourism and heritage trails; 23 new or improved bridges or crossings; 22 new signage points; installation of public art; 13 ha of upgraded parks; 2 multi-use games areas; 2 new toilets. | Supportive active transport infrastructure (e.g. cycleways and traffic calming) | MVPA Physical activity |
| Jones, 2012^102^ | **Environmental restructuring (physical)** | Section of a traffic-free cycle route. | Supportive active transport infrastructure (e.g. cycleways and traffic calming) | Cycling frequency |
| Karpinski, 2021^104^ | **Environmental restructuring (physical)** | Installation of a protected bike lane in Boston, Massachusetts. | Supportive active transport infrastructure (e.g. cycleways and traffic calming) | Cycling frequency |
| Keall, 2015^106^ | **Environmental restructuring (physical),** Environmental restructuring (social) | The Model Communities Programme: public investment in infrastructure and programmes to encourage active travel. | Supportive active transport infrastructure (e.g. cycleways and traffic calming) | AT frequency |
| Keall, 2022^105^ | **Environmental restructuring (physical),**  Environmental restructuring (social) | Funding to install walking and cycling infrastructure and run programs to promote and normalise active travel | Supportive active transport infrastructure (e.g. cycleways and traffic calming) | AT frequency |
| Lambe, 2017^110^ | Education,  **Environmental restructuring (physical),** Environmental restructuring (social) | Town 1: developed 12 km of orbital pedestrian/cycleways, improved existing cycleways on the town’s radial routes, improved the public realm and pedestrian infrastructure in the town centre, created 1.6 km river boardwalks and organized three Active Travel themed campaigns. Active travel promotional materials were sent home to the parents of all primary school children encouraging them to allow their children to walk or cycle to school and to model the behaviour themselves.   Town 2: infrastructural measures to create safer routes to four of the town’s five primary schools. | Supportive active transport infrastructure (e.g. cycleways and traffic calming) | Walking frequency Cycling frequency Modal shift frequency |
| Limb, 2020^111^ | **Environmental restructuring (physical)** | A purpose-built mixed-use residential development built on active design principles to encourage active living by improving neighbourhood walkability, access to public transport and open space with restrictions on vehicle parking. | Supportive active transport infrastructure (e.g. cycleways and traffic calming) | Walking duration Cycling duration AT frequency Modal shift duration |
| McDonald, 2014^116^ | **Environmental restructuring (physical),** Environmental restructuring (social) | Safe Routes to School: provided grants to assist communities in creating safer and more supportive environments for children to walk or bicycle to school. | Supportive active transport infrastructure (e.g. cycleways and traffic calming) | AT frequency |
| Mitra, 2021^124^ | **Environmental restructuring (physical)** | Cycling facilities (cycle track, painted or buffered bicycle lane). | Supportive active transport infrastructure (e.g. cycleways and traffic calming) | Cycling frequency |
| Ostergaard, 2015^127^ | Incentivisation,  Training,  **Environmental restructuring (physical**) | Structural changes near schools e.g. road surface, signposting and traffic regulation such as one-way streets and regulation of car drop off zones and educational and motivational interventions e.g. competitions and monitoring and cycling safety. | Supportive active transport infrastructure (e.g. cycleways and traffic calming) | Cycling frequency |
| Rissel, 2015^155^ | **Environmental restructuring (physical)** | Construction of a new bicycle path (2.4 km bi-directional separated bicycle path in inner-Sydney). | Supportive active transport infrastructure (e.g. cycleways and traffic calming) | Cycling frequency |
| Rothman, 2022^133^ | **Environmental restructuring (physical)** | Vision Zero school safety zone interventions. Included speed feedback signs, flashing beacons, ‘school’ and speed limit markings and other pavement markings (e.g., zebra bars, crosswalk lines). | Supportive active transport infrastructure (e.g. cycleways and traffic calming) | AT frequency |
| Stappers, 2021^143^ | **Environmental restructuring (physical)** | The Green Carpet. Two one-way streets separated by a semi-paved middle section, prioritised for use by pedestrians, cyclists and for recreation. | Supportive active transport infrastructure (e.g. cycleways and traffic calming) | Physical activity MVPA |
| Sun, 2020^145^ | **Environmental restructuring (physical)** | New train station. | New public transport service (e.g. new train line or light rail service) | Walking frequency Walking duration Cycling frequency Cycling duration Modal shift duration |
| Xiao, 2022^152^ | **Environmental restructuring (physical)** | 15 new or improved cycle lanes in Paris and Lyon. | Supportive active transport infrastructure (e.g. cycleways and traffic calming) | Cycling frequency |
| Gu, 2019^90^ | **Environmental restructuring (physical)** | New public transport system. | New public transport service (e.g. new train line or light rail service) | Cycling frequency |
| He, 2022^92^ | **Environmental restructuring (physical)** | New metro rail line. | New public transport service (e.g. new train line or light rail service) | Physical activity |
| Hong, 2016^97^ | **Environmental restructuring (physical)** | New light rail transit service. | New public transport service (e.g. new train line or light rail service) | Walking frequency Modal shift frequency |
| Spears, 2016^142^ | **Environmental restructuring (physical)** | New light rail transit service. | New public transport service (e.g. new train line or light rail service) | Walking frequency Cycling frequency Modal shift frequency |
| Olsen, 2016^126^ | **Environmental restructuring (physical)** | New motorway infrastructure, M74 motorway extension. | Infrastructure for motorised transport | AT frequency |
| Aittasalo, 2012 & 2019^49, 50^ | Education, Incentivisation, **Environmental restructuring (social),** Enablement | Workplace-specific social and behavioural strategies. One group meeting, monitored pedometer-use and six e-mail messages. | Workplace-specific educational programme | Walking duration Walking frequency  AT frequency Modal shift duration Modal shift frequency |
| Brownson, 2004^64^ | **Environmental restructuring (social)** | Developed with community input and included individually tailored newsletters, interpersonal activities that stressed social support, and community-wide events such as walk-a-thons. | Community developed interventions to target physical inactivity (e.g. community “fun-walks”) | Walking frequency Walking duration |
| Jancey, 2011^101^ | Persuasion,  **Environmental restructuring (social)** | Walking program where trained walk leaders supervised walking groups twice weekly. Participants were provided with a progressive weekly exercise program containing written info on walking. Home-based booster involved an interactive booklet. | Community walking programme | Walking duration Walking frequency |
| Merom, 2003^122^ | **Environmental restructuring (social),** Environmental restructuring (physical) | Local promotional campaign around a newly constructed rail trail in western Sydney, Australia. | Promotion of new walking infrastructure | Walking duration AT frequency |
| Kubota, 2020^108^ | Education,  **Environmental restructuring (social)** | Monthly walking maps and walking events. | Provision of walking maps and events | Walking duration Walking frequency |
| Rissel, 2010^131^ | Education,  Persuasion,  Training,  **Environmental restructuring (social)** | The Cycling Connecting Communities (CCC) Project: a community-based cycling promotion program that included a range of community engagement and social marketing activities, such as organised bike rides and events, cycling skills courses, the distribution of cycling maps of the area and coverage in the local press. | Community based cycling promotion program | Cycling duration Cycling frequency |
| Lambe, 2022^109^ | Incentivisation,  **Environmental restructuring (social)** | Beat the Street (BTS) game: Beat Boxes (sensors installed on lampposts) for children were installed in 48 locations, fob keys provided to children and BTS cards to parents, location maps and information letters were distributed to every child attending school in the area. Playing the game consisted of tapping the card or fob on at least 2 Beat Boxes within an hour to earn points. | Gamification of walking to school | AT frequency |
| Coombes, 2016^72^ | Incentivisation,  **Environmental restructuring (social)** | 40 Beat Boxes (sensors installed on lampposts) were installed in the street environment. Participants were awarded a point each time they touched their smartcard on a sensor, allowing children to compete against to see who could achieve the most points. | Gamification of walking to school | MVPA |
| Petrunoff, 2016^128^ | Incentivisation,  Environmental restructuring (physical), **Environmental restructuring (social)** | Policy changes for hospital travel policy, (e.g. completing a parking management study to inform a new parking policy, public transport ticket salary deduction/sacrifice schemes), and new infrastructure (e.g. provision of end of trip facilities, marking of carpooling spaces in staff car park) and behaviour change (e.g. cycling and walking programs). | Hospital based active transport campaign | AT frequency, Modal shift |
| Christiansen, 2014^68^ | Environmental restructuring (physical), **Environmental restructuring (social)** | Comprehensive school-based intervention to improve non-curricular physical activity through changes of the physical and organisational environment supported by educational activities. Policy, program and physical initiatives (a short cycle path near the school separating bike riders from cars, speed humps, a new parking area further from the school and 30 cycles to use for educational purposes). | School based active transport campaign | Walking frequency Cycling frequency AT frequency Modal shift frequency |
| Elinder, 2012^80^ | Persuasion,  **Environmental restructuring (social)** | Tailored action plans developed by school health teams on the basis of a self-assessment questionnaire assessing strengths and weaknesses of each school’s health practices and environments. | Tailored active transport action plans for schools | AT frequency |
| Gutierrez, 2014^91^ | Environmental restructuring (physical), **Environmental restructuring (social)** | Positioning of the newly hired crossing guards and the implementation of an awareness campaign | New crossing guards and road safety awareness campaign | AT frequency |
| Malakellis, 2017^114^ | Environmental restructuring (physical), **Environmental restructuring (social),** Enablement | Intervention schools were provided $50k AUD funding to cover costs relating redeveloping the school environment to support nutrition and physical activity, sporting/fitness equipment, and presentations health promotion from community leaders. | School based physical activity campaign | AT frequency |
| McKee, 2007^117^ | **Environmental restructuring (social)** | Travelling Green: a school-based active travel project. Interactive resources provided to classroom teachers, children and their families including customised map, educational resources, goal-setting activities and safety accessories. | School based active transport campaign | AT frequency Modal shift distance |
| McMinn, 2012^118^ | **Environmental restructuring (social)** | Travelling Green: a school-based active travel project. The teacher's handbook contains introductory activities to the intervention and 13 lesson plans on topics such as road safety, the importance of a healthy lifestyle, and how the body functions. The pupil pack contains materials that encourage the pupils to set walking goals and to record how they travel to and from school each day. | School based active transport campaign | MVPA |
| Jordan, 2008^103^ | **Environmental restructuring (social)** | Gold Medal Schools are encouraged to promote fruits and vegetables at breakfast and lunch and to participate in physical activity programs such as Walk Your Child to School Day and the President's Challenge for physical fitness. | School based active transport campaign | Physical activity |
| Sahlqvist, 2019^135^ | Education,  Incentivisation,  **Environmental restructuring (social)** | Walk to School campaign: participating schools received campaign materials including posters and classroom calendars. Schools were encouraged to host their own activities, such as competitions and one-day promotional events. VicHealth also engaged in wider promotion of the campaign to parents and the broader community through online advertising, marketing, public relations activities and social media. | School based walk to school campaign | AT frequency |
| Simon, 2008 & 2014^138, 139^ | Education,  **Environmental restructuring (social)** | School based intervention: educational component focusing on physical activity and sedentary behaviours. Opportunities for physical activity were offered at lunchtimes, breaks and afterschool hours. Sporting events and ‘cycling to school’ days were organized. Parents and educators were encouraged to provide support to enhance the adolescents’ physical activity level. | School based physical activity campaign | AT frequency  Physical activity |
| Xu, 2015^153^ | Education,  **Environmental restructuring (social)** | One-year multi-component intervention program (classroom curriculum, school environment support, family involvement and fun programs/events) together with routine health education. | School based promotional campaign | AT frequency |
| Buckley, 2013^65^ | **Environmental restructuring (social)** | Safe Routes to School (SR2S) program. Three designated days for walking and bicycling; one in fall, one in winter and one in spring. | Designated ride and walk to school days | Walking frequency Cycling frequency |
| Bungum, 2014^66^ | **Environmental restructuring (social)** | Nevada Moves Day: a state-wide celebration of the Safe Routes to School Program that encourages kids and their families to walk or bicycle to and from school. Promoted at intervention schools with morning announcements and sheet reminders. | Designated ride and walk to school days | AT frequency Motor vehicle counts Modal shift frequency |
| Mendoza, 2017^119^ | **Environmental restructuring (social),** Enablement | Bicycle train offered daily (i.e., students volunteered to cycle with study staff to and from school). | School based bicycle train | Cycling frequency |
| Folta, 2013^83^ | Education,  Incentivisation,  **Environmental restructuring (social)** | Walk to School Campaign: walking school bus, traffic calming, walking contest, maps for safe routes. | School based walking bus | Walking frequency |
| Heelan, 2009^93^ | **Environmental restructuring (social)** | Walking school bus with neighbourhood walk-stops designated within a 1-mile radius of the intervention schools. | School based walking bus | AT frequency |
| Mendoza, 2009 & 2011^120, 121^ | **Environmental restructuring (social)** | Walking school bus program consisting of a part-time WSB coordinator and parent volunteers. | School based walking bus | AT frequency |
| Rowland, 2003^134^ | Persuasion,  **Environmental restructuring (social)** | Site specific advice from a school travel coordinator on school travel patterns. | School based walking bus | Walking frequency AT frequency Modal shift |
| Sirard, 2008^141^ | **Environmental restructuring (social)** | Walking school bus run by school volunteers. | School based walking bus | MVPA |
| Bernstein, 2017^58^ | Persuasion, Training, **Enablement** | Provision of a bicycle and ten group sessions over 12-weeks consisting of on-road education, group rides and bicycle safety classroom instruction. | Provision of a bicycle and on-road training and education | Cycling frequency |
| Andersson, 2021^53^ | **Enablement** | Provision of e-bikes to frequent motor vehicle drivers in Sweden. | Provision of e-bikes | Walking frequency Cycling frequency Walking distance Cycling distance Modal shift frequency |
| Bjornara, 2019^60^ | **Enablement** | Three months' access to an e-bike with trailer, longtail bike or traditional bike with trailer. | Provision of bikes or e-bikes | Cycling duration Cycling distance Modal shift frequency |
| Fyhri, 2015^85^ | **Enablement** | Provision of e-bikes on a trial period to individuals who took part in a travel behaviour survey in Norway. | Provision of e-bikes | Cycling frequency |
| Hosford, 2019^98^ | **Enablement** | Newly implemented and existing public bicycle share programs. | Bicycle share program | Cycling frequency Cycling duration |
| Grimes, 2020^88^ | **Enablement** | Free 1-month membership to Kansas City’s bike share system, B-cycle. | Free bike share membership | Walking frequency Walking duration, Cycling frequency Cycling duration Step count |
| Brown, 2003^63^ | **Enablement** | BruinGO: Fare-free transit to members of the university community at the University of California, Los Angeles. | Fare-free public transport | Walking frequency Cycling frequency Modal shift frequency |
