## Supplementary material for "Effectiveness of interventions for modal shift to walking and bike riding: a systematic review with meta-analysis": Data Supplement 4

Data Supplement 4 – Quality Appraisal

Table 1. Quality Appraisal Results of Randomised Controlled Trials

| First Author, Year Published | True randomisation of participants to treatment groups | Treatment group allocation concealment | Treatment groups similar at baseline | Participants blinded to treatment assignment | Individuals delivering treatment blind to participant assignment | Identical treatment other than intervention difference | Outcome assessors blind to treatment assignment | Outcome measurement identical in all treatment groups | Outcomes measured reliably | Follow-up conducted, or difference between groups described | Participants analysed within group they were randomized to | Appropriate statistical analysis | Trial design appropriate |
| --- | --- | --- | --- | --- | --- | --- | --- | --- | --- | --- | --- | --- | --- |
| Aittasalo, 2019 | ✕ | ✕ | ✓ | ✕ | ✕ | ✓ | ✕ | ✓ | ✓ | ✕ | ✕ | ✓ | ✓ |
| Aittasalo, 2012 | ✓ | ✕ | ✓ | ✕ | ✕ | ✓ | ✕ | ✓ | ✓ | ✕ | ✕ | ✓ | ✓ |
| Andersson, 2021 | ✕ | ✕ | ✓ | ✕ | ✕ | ✓ | ✕ | ✓ | ✓ | ✓ | ✕ | ✕ | ✓ |
| Aranda-Balboa, 2022 | ✕ | ✕ | ✕ | ✕ | ✕ | ✓ | ✕ | ✓ | ✓ | ✓ | ✓ | ✓ | ✓ |
| Audrey, 2019 | ✓ | ✓ | ✓ | ✕ | ✕ | ✓ | ✕ | ✓ | ✓ | ✓ | ✓ | ✓ | ✓ |
| Bernstein, 2017 | ✓ | ✓ | ✕ | ✕ | ✕ | ✓ | ✕ | ✓ | ✓ | ✕ | ✓ | ✓ | ✓ |
| Bjornara, 2019 | ✕ | ✕ | ✓ | ✕ | ✕ | ✓ | ✕ | ✓ | ✓ | ✓ | ✕ | ✓ | ✓ |
| Christiansen, 2014 | ✕ | ✕ | ✓ | ✕ | ✕ | ✓ | ✕ | ✓ | ✓ | ✓ | ✓ | ✓ | ✓ |
| Ciccone, 2021 | ✕ | ✕ | ✓ | ✕ | ✕ | ✓ | ✕ | ✓ | ✓ | ✓ | ✕ | ✓ | ✓ |
| Coffeng, 2014 | ✓ | ✕ | ✓ | ✕ | ✕ | ✓ | ✕ | ✓ | ✓ | ✓ | ✓ | ✓ | ✓ |
| Ducheyne, 2014 | ✕ | ✕ | ✕ | ✕ | ✕ | ✓ | ✕ | ✓ | ✓ | ✕ | ✕ | ✓ | ✓ |
| Duncan, 2011 | ✕ | ✕ | ✕ | ✕ | ✕ | ✕ | ✕ | ✓ | ✓ | ✓ | ✕ | ✓ | ✓ |
| Dunton, 2008 | ✓ | ✕ | ✓ | ✕ | ✕ | ✓ | ✕ | ✓ | ✓ | ✓ | ✓ | ✓ | ✓ |
| Ek, 2020 | ✓ | ✓ | ✓ | ✕ | ✕ | ✓ | ✓ | ✓ | ✓ | ✓ | ✓ | ✓ | ✓ |
| Fyhri, 2015 | ✕ | ✕ | ✕ | ✕ | ✕ | ✓ | ✕ | ✓ | ✓ | ✓ | ✕ | ✓ | ✓ |
| Grimes, 2020 | ✓ | ✓ | ✕ | ✕ | ✕ | ✓ | ✕ | ✓ | ✓ | ✕ | ✕ | ✓ | ✓ |
| Isensee, 2018 | ✓ | ✓ | ✕ | ✕ | ✕ | ✓ | ✕ | ✓ | ✓ | ✓ | ✓ | ✓ | ✓ |
| King, 2017 | ✓ | ✓ | ✓ | ✓ | ✕ | ✓ | ✕ | ✓ | ✓ | ✕ | ✓ | ✓ | ✓ |
| Liddle, 2014 | ✓ | ✓ | ✓ | ✕ | ✕ | ✓ | ✓ | ✓ | ✓ | ✓ | ✓ | ✓ | ✓ |
| Maca, 2020 | ✕ | ✕ | ✕ | ✕ | ✕ | ✓ | ✕ | ✓ | ✓ | ✕ | ✕ | ✓ | ✓ |
| Mackey, 2019 | ✓ | ✕ | ✓ | ✕ | ✕ | ✓ | ✓ | ✓ | ✓ | ✓ | ✓ | ✓ | ✓ |
| Mendoza, 2017 | ✓ | ✕ | ✕ | ✕ | ✕ | ✓ | ✕ | ✓ | ✓ | ✓ | ✓ | ✓ | ✓ |
| Mendoza, 2011 | ✓ | ✕ | ✕ | ✕ | ✕ | ✓ | ✕ | ✓ | ✓ | ✓ | ✓ | ✓ | ✓ |
| Merom, 2007 | ✓ | ✓ | ✓ | ✕ | ✕ | ✓ | ✓ | ✓ | ✓ | ✓ | ✓ | ✓ | ✓ |
| Mutrie, 2002 | ✓ | ✓ | ✕ | ✕ | ✕ | ✓ | ✕ | ✓ | ✓ | ✓ | ✓ | ✓ | ✓ |
| Purath, 2005 | ✕ | ✕ | ✕ | ✕ | ✕ | ✓ | ✕ | ✓ | ✓ | ✕ | ✕ | ✓ | ✓ |
| Purath, 2004 | ✕ | ✕ | ✕ | ✕ | ✕ | ✓ | ✕ | ✓ | ✓ | ✓ | ✕ | ✓ | ✓ |
| Rowland, 2003 | ✓ | ✓ | ✓ | ✕ | ✕ | ✓ | ✕ | ✓ | ✓ | ✓ | ✓ | ✓ | ✕ |
| Salinas, 2019 | ✕ | ✕ | ✕ | ✓ | ✕ | ✓ | ✕ | ✓ | ✓ | ✕ | ✓ | ✓ | ✓ |
| Simon, 2014 | ✕ | ✕ | ✕ | ✕ | ✕ | ✓ | ✕ | ✓ | ✓ | ✓ | ✓ | ✓ | ✓ |
| Simon, 2008 | ✕ | ✕ | ✕ | ✕ | ✕ | ✓ | ✕ | ✓ | ✓ | ✓ | ✓ | ✓ | ✓ |
| Singh, 2009 | ✓ | ✕ | ✕ | ✕ | ✕ | ✓ | ✕ | ✓ | ✓ | ✓ | ✓ | ✓ | ✓ |
| Sirard, 2008 | ✕ | ✕ | ✕ | ✕ | ✕ | ✓ | ✕ | ✓ | ✓ | ✓ | ✕ | ✓ | ✓ |
| Suchert, 2015 | ✓ | ✕ | ✕ | ✕ | ✕ | ✓ | ✕ | ✓ | ✓ | ✓ | ✓ | ✓ | ✓ |
| Van Dyck, 2016 | ✓ | ✓ | ✕ | ✕ | ✕ | ✓ | ✕ | ✓ | ✓ | ✓ | ✕ | ✓ | ✓ |
| van Stralen, 2010 | ✕ | ✕ | ✕ | ✕ | ✕ | ✓ | ✕ | ✓ | ✓ | ✓ | ✕ | ✓ | ✕ |
| Xu, 2015 | ✓ | ✓ | ✓ | ✕ | ✕ | ✓ | ✕ | ✓ | ✓ | ✓ | ✕ | ✓ | ✕ |

Table 2. Quality Appraisal of Quasi-Experimental Studies and Interrupted Time Series

| First author, year published | Cause and effect definitions clear | Participants in comparison group were similar | Comparison group received similar treatment other than intervention | Control group identified | Multiple measurements of outcome taken pre and post intervention | Follow up conducted or differenced in follow up described and analysed | Outcome measurement identical in all treatment groups | Outcomes measured reliably | Appropriate statistical analysis |
| --- | --- | --- | --- | --- | --- | --- | --- | --- | --- |
| Aldred, 2021 | ✓ | ✓ | ✓ | ✓ | ✓ | ✕ | ✓ | ✓ | ✕ |
| Aldred, 2019 | ✓ | ✓ | ✓ | ✓ | ✕ | ✕ | ✓ | ✓ | ✕ |
| Auchincloss, 2019 | ✓ | ✓ | ✓ | ✓ | ✓ | ✕ | ✓ | ✓ | ✓ |
| Benton, 2021 | ✓ | ✓ | ✓ | ✓ | ✓ | ✕ | ✓ | ✓ | ✓ |
| Biondi, 2022 | ✓ | ✕ | ✓ | ✓ | ✓ | ✕ | ✓ | ✓ | ✓ |
| Boarnet, 2005 | ✓ | ✕ | ✕ | ✓ | ✓ | ✕ | ✓ | ✓ | ✓ |
| Brown, 2016 | ✓ | ✕ | ✓ | ✓ | ✓ | ✕ | ✓ | ✓ | ✓ |
| Brown, 2003 | ✓ | ✕ | ✓ | ✓ | ✓ | ✕ | ✓ | ✓ | ✓ |
| Brownson, 2004 | ✓ | ✓ | ✓ | ✓ | ✓ | ✕ | ✓ | ✓ | ✓ |
| Buckley, 2013 | ✓ | ✕ | ✓ | ✓ | ✓ | ✕ | ✓ | ✓ | ✓ |
| Bungum, 2014 | ✓ | ✓ | ✓ | ✓ | ✓ | ✕ | ✓ | ✓ | ✓ |
| Cambra, 2020 | ✓ | ✕ | ✓ | ✓ | ✓ | ✕ | ✓ | ✓ | ✓ |
| Clark, 2014 | ✓ | ✓ | ✓ | ✓ | ✓ | ✕ | ✓ | ✓ | ✓ |
| Coombes, 2016 | ✓ | ✓ | ✓ | ✓ | ✓ | ✓ | ✓ | ✓ | ✓ |
| Crane, 2017 | ✓ | ✕ | ✓ | ✓ | ✓ | ✕ | ✓ | ✓ | ✓ |
| Crawford, 2013 | ✓ | ✕ | ✓ | ✓ | ✓ | ✕ | ✓ | ✓ | ✓ |
| Dill, 2014 | ✓ | ✓ | ✓ | ✓ | ✓ | ✕ | ✓ | ✓ | ✓ |
| Elinder, 2012 | ✓ | ✓ | ✓ | ✓ | ✓ | ✓ | ✓ | ✓ | ✓ |
| Felix, 2020 | ✓ | ✕ | ✓ | ✓ | ✓ | ✕ | ✓ | ✓ | ✓ |
| Fitzhugh, 2010 | ✓ | ✕ | ✓ | ✓ | ✓ | ✕ | ✓ | ✓ | ✓ |
| Folta, 2013 | ✓ | ✕ | ✓ | ✓ | ✓ | ✓ | ✓ | ✓ | ✓ |
| Frank, 2021 | ✓ | ✕ | ✓ | ✓ | ✓ | ✕ | ✓ | ✓ | ✓ |
| Fuller, 2012 | ✓ | ✕ | ✓ | ✓ | ✓ | ✕ | ✓ | ✓ | ✓ |
| Fuller, 2019 | ✓ | Not applicable | Not applicable | Not applicable | ✓ | ✕ | ✓ | ✓ | ✓ |
| Geng, 2020 | ✓ | ✓ | ✓ | ✓ | ✓ | ✕ | ✓ | ✓ | ✓ |
| Goodman, 2013 | ✓ | ✓ | ✓ | ✓ | ✓ | ✕ | ✓ | ✓ | ✓ |
| Goodman, 2016 | ✓ | ✕ | ✓ | ✓ | ✓ | ✓ | ✓ | ✓ | ✓ |
| Grunseit, 2019 | ✓ | Not applicable | Not applicable | Not applicable | ✓ | ✕ | ✓ | ✓ | ✓ |
| Gu, 2019 | ✓ | ✕ | ✓ | ✓ | ✓ | ✕ | ✓ | ✓ | ✓ |
| Gutierrez, 2014 | ✓ | ✓ | ✓ | ✓ | ✓ | ✕ | ✓ | ✓ | ✓ |
| He, 2022 | ✓ | ✓ | ✓ | ✓ | ✓ | ✕ | ✓ | ✓ | ✓ |
| Heelan, 2009 | ✓ | ✓ | ✓ | ✓ | ✓ | ✕ | ✓ | ✓ | ✓ |
| Heesch, 2016 | ✓ | ✕ | ✓ | ✓ | ✓ | ✓ | ✓ | ✓ | ✓ |
| Heinrich, 2011 | ✓ | ✕ | ✓ | ✓ | ✓ | ✕ | ✓ | ✓ | ✕ |
| Hoelscher, 2016 | ✓ | ✓ | ✓ | ✓ | ✓ | ✕ | ✓ | ✓ | ✓ |
| Hong, 2016 | ✓ | ✓ | ✓ | ✓ | ✓ | ✓ | ✓ | ✓ | ✓ |
| Hosford, 2019 | ✓ | ✓ | ✓ | ✓ | ✓ | ✕ | ✓ | ✓ | ✓ |
| Hunter, 2021 | ✓ | ✕ | ✓ | ✓ | ✓ | ✓ | ✓ | ✓ | ✓ |
| Jancey, 2011 | ✓ | ✓ | ✓ | ✓ | ✓ | ✓ | ✓ | ✓ | ✓ |
| Jones, 2012 | ✓ | ✓ | ✓ | ✓ | ✕ | ✕ | ✓ | ✓ | ✓ |
| Jordan, 2008 | ✓ | ✓ | ✓ | ✓ | ✓ | ✕ | ✓ | ✓ | ✓ |
| Karpinski, 2021 | ✓ | ✕ | ✓ | ✓ | ✓ | ✕ | ✓ | ✓ | ✓ |
| Keall, 2022 | ✓ | ✓ | ✓ | ✓ | ✓ | ✕ | ✓ | ✓ | ✕ |
| Keall, 2015 | ✓ | ✓ | ✓ | ✓ | ✓ | ✕ | ✓ | ✓ | ✓ |
| Kubota, 2020 | ✓ | ✕ | ✓ | ✓ | ✓ | ✓ | ✓ | ✓ | ✓ |
| Lambe, 2022 | ✓ | ✕ | ✕ | ✓ | ✓ | ✕ | ✓ | ✓ | ✓ |
| Lambe, 2017 | ✓ | ✕ | ✓ | ✓ | ✓ | ✕ | ✓ | ✓ | ✓ |
| Limb, 2020 | ✓ | ✕ | ✓ | ✓ | ✓ | ✕ | ✓ | ✓ | ✓ |
| Malakellis, 2017 | ✓ | ✕ | ✓ | ✓ | ✓ | ✓ | ✓ | ✓ | ✓ |
| Mayer, 1982 | ✓ | ✕ | ✓ | ✓ | ✓ | ✕ | ✓ | ✓ | ✓ |
| McDonald, 2014 | ✓ | ✕ | ✓ | ✓ | ✓ | ✕ | ✓ | ✓ | ✓ |
| McKee, 2007 | ✓ | ✕ | ✓ | ✓ | ✓ | ✕ | ✓ | ✓ | ✓ |
| McMinn, 2012 | ✓ | ✕ | ✓ | ✓ | ✓ | ✕ | ✓ | ✓ | ✓ |
| Mendoza, 2009 | ✓ | ✓ | ✓ | ✓ | ✓ | ✕ | ✓ | ✓ | ✓ |
| Merom, 2003 | ✓ | ✕ | ✓ | ✓ | ✓ | ✕ | ✓ | ✓ | ✓ |
| Mitra, 2021 | ✓ | ✓ | ✓ | ✓ | ✕ | ✕ | ✓ | ✓ | ✓ |
| Olsen, 2016 | ✓ | ✓ | ✓ | ✓ | ✓ | ✕ | ✓ | ✓ | ✓ |
| Ostergaard, 2015 | ✓ | ✕ | ✓ | ✓ | ✓ | ✕ | ✓ | ✓ | ✓ |
| Petrunoff, 2016 | ✓ | Not applicable | Not applicable | Not applicable | ✓ | ✕ | ✓ | ✓ | ✓ |
| Rissel, 2010 | ✓ | ✓ | ✓ | ✓ | ✓ | ✓ | ✓ | ✓ | ✓ |
| Rissel, 2015 | ✓ | ✕ | ✓ | ✓ | ✓ | ✓ | ✓ | ✓ | ✓ |
| Rothman, 2022 | ✓ | ✓ | ✓ | ✓ | ✓ | ✕ | ✓ | ✓ | ✓ |
| Sahlqvist, 2019 | ✓ | ✕ | ✕ | ✓ | ✓ | ✕ | ✓ | ✓ | ✓ |
| Sersli, 2019 | ✓ | ✕ | ✓ | ✓ | ✓ | ✕ | ✓ | ✓ | ✓ |
| Spears, 2016 | ✓ | ✓ | ✓ | ✓ | ✓ | ✓ | ✓ | ✓ | ✓ |
| Stappers, 2021 | ✓ | ✕ | ✓ | ✓ | ✓ | ✓ | ✓ | ✓ | ✓ |
| Sun, 2020 | ✓ | ✕ | ✓ | ✓ | ✓ | ✓ | ✓ | ✓ | ✓ |
| Sundfor, 2022 | ✓ | ✕ | ✓ | ✓ | ✓ | ✕ | ✓ | ✓ | ✓ |
| Villa-Gonzalez, 2017 | ✓ | ✕ | ✓ | ✓ | ✓ | ✕ | ✓ | ✓ | ✓ |
| Villa-Gonzalez, 2016 | ✓ | ✓ | ✓ | ✓ | ✓ | ✕ | ✓ | ✓ | ✓ |
| Walsh, 2021 | ✓ | ✕ | ✓ | ✓ | ✓ | ✕ | ✓ | ✓ | ✓ |
| Xiao, 2022 | ✓ | ✕ | ✕ | ✓ | ✓ | ✕ | ✓ | ✓ | ✓ |

Table 3. Extended description of quality assessment for randomised controlled trials

| First Author, Year Published | True randomisation of participants to treatment groups | Treatment group allocation concealment | Treatment groups similar at baseline | Participants blinded to treatment assignment | Individuals delivering treatment blind to participant assignment | Identical treatment other than intervention difference | Outcome assessors blind to treatment assignment | Outcome measurement identical in all treatment groups | Outcomes measured reliably | Follow-up conducted, or difference between groups described | Participants analysed within group they were randomized to | Appropriate statistical analysis | Trial design appropriate |
| --- | --- | --- | --- | --- | --- | --- | --- | --- | --- | --- | --- | --- | --- |
| Aittasalo, 2019 | ✕ Cluster randomisation was conducted. Insufficient information was provided about the sequence generation process. | ✕ No mention+F15 | ✓ | ✕ No mention of blinding. Unlikely to have been possible due to intervention type | ✕ No mention of blinding. Unlikely to have been possible due to intervention type | ✓ | ✕ No mention of blinding. Unlikely to have occurred. | ✓ | ✓ Reliance on self-reported information (survey). Objective data not analysed as very few participants used accelerometer and travel diary. | ✕ Only self-reported ACW were analysed as not enough objective data was collected. High   accelerometer dropout rate. | ✕ ITT analysis not reported | ✓ Statistical power was calculated and achieved. Mann-Whitney U-test used and seemed appropriate. | ✓ Cluster RCT. Intra-cluster correlation reported. |
| Aittasalo, 2012 | ✓ Used computer-generated randomisation lists (stratified randomisation with random allocation sequences) | ✕ No mention | ✓ | ✕ No mention | ✕ No mention | ✓ | ✕ No mention | ✓ Both groups completed follow-up questionnaires at 2, 6 and 12 months | ✓ Self-reported outcomes (survey). Objective measures (logbook and pedometer) were used as an intervention not an outcome measure. | ✕ Higher FU in intervention group. | ✕ ITT analysis not reported | ✓ | ✓ Individual-level randomisation |
| Andersson, 2021 | ✕ Not adequately described | ✕ No mention | ✓ | ✕ No mention | ✕ No mention | ✓ | ✕ No mention | ✓ | ✓ GPS-tracker app used to collect data. Included a mixture of self-reported and objective data. | ✓ High LTFU, especially in control group. Droupout analysis conducted which showed that FU participants had no statistically significant difference in demographics compared to the other participants. | ✕ ITT analysis not reported | ✕ Small sample size limits statistical power. | ✓ |
| Aranda-Balboa, 2022 | ✕ Randomisation process briefly described, insufficient information regarding sequence generation | ✕ No mention | ✕ Significant difference in bike ownership and active commute to school at baseline | ✕ No mention | ✕ No mention | ✓ | ✕ No mention | ✓ | ✓ Accelerometer, activity diary, GPS unit and family questionnaire | ✓ | ✓ ITT analysis conducted | ✓ Sufficient sample size for statistical power | ✓ |
| Audrey, 2019 | ✓ Randomly computer generated allocation done by an external statistician | ✓ | ✓ | ✕ Given the nature of the intervention it was not possible to blind participants following randomisation. | ✕ Unable to blind due to nature of intervention | ✓ | ✕ No mention | ✓ | ✓ Accelerometer, GPS data and travel diary | ✓ 3 workplaces dropped out in intervention group, reasons for dropout given. No dropout in control. Adequate analysis. | ✓ ITT analysis conducted (mentioned in protocol) | ✓ | ✓ Cluster RCT. Intra-cluster correlation reported. |
| Bernstein, 2017 | ✓ Participants were randomized 1:1 to the intervention or control groups, stratified by site. Random assignment made using REDCap | ✓ It was not possible to determine group assignment prior to randomizing each individual. | ✕ Significant difference in income | ✕ Non-blinded | ✕ Non-blinded | ✓ | ✕ Non-blinded | ✓ | ✓ Survey only | ✕ Similar level of LTFU for both groups, not described in much detail | ✓ ITT analysis conducted | ✓ | ✓ Pilot RCT to determine feasibility |
| Bjornara, 2019 | ✕ Not adequately described | ✕ No mention | ✓ | ✕ No mention | ✕ No mention | ✓ | ✕ No mention | ✓ | ✓ Survey only | ✓ 1 dropout in control group due to illness | ✕ ITT analysis not reported | ✓ | ✓ |
| Christiansen, 2014 | ✕ Cluster randomisation w/ matched pair design. Lack of info on true randomisation process. | ✕ No mention | ✓ | ✕ No mention | ✕ Unable to blind due to nature of intervention | ✓ | ✕ No mention | ✓ | ✓ Travel diary and questionnaire | ✓ Slightly higher LTFU in control, reasons for LTFU adequately described. | ✓ ITT analysis conducted | ✓ Multilevel logistic regression used | ✓ Cluster RCT. Intraclass/intracluster correlation reported. |
| Ciccone, 2021 | ✕ Stratification used but randomisation process not adequately described | ✕ No mention | ✓ No significant differences in characteristics at baseline. Shown in Appendix B, Table 6 | ✕ No mention | ✕ No mention | ✓ | ✕ No mention | ✓ | ✓ App registers travel behaviour automatically w 82% accuracy, has an option for manual route correction | ✓ Dropouts reported with attrition rates calculated for both groups. | ✕ ITT analysis not reported | ✓ | ✓ |
| Coffeng, 2014 | ✓ Randomisation executed by an independent researcher, by using a computer-generated list from SPSS | ✕ No mention | ✓ | ✕ Unable to blind due to nature of intervention | ✕ Unable to blind due to nature of intervention | ✓ | ✕ No mention | ✓ | ✓ Survey | ✓ Provided reasons for LTFU, reason unknown for 25 participants. Wasn't clear abt which groups sustained LTFU, an overall number was given instead | ✓ ITT analysis conducted | ✓ Linear mixed model analysis used | ✓ 2x2 factorial design. Intra-class coefficient calculated |
| Ducheyne, 2014 | ✕ Not adequately described | ✕ No mention | ✕ Difference in normal weight and SES | ✕ No mention | ✕ Not blinded | ✓ | ✕ No mention | ✓ | ✓ Practical cycling test and parent questionnaire | ✕ LTFU not adequately reported | ✕ ITT analysis not reported | ✓? No mention of power | ✓ Cluster RCT? Intra-cluster correlation reported. |
| Duncan, 2011 | ✕ Not adequately described | ✕ No mention | ✕ Baseline demographics not reported adequately. Statistically sig diff for unhealthy drink consumption at baseline | ✕ No mention | ✕ No mention | ✕ It is probable that a certain amount of class contamination occurred, such that the behaviour of the control participants was affected by the experiences of the intervention participants as they progressed through the programme. | ✕ No mention | ✓ | ✓ Pedometer and diary | ✓ | ✕ ITT analysis not reported | ✓ | ✓ |
| Dunton, 2008 | ✓ Random allocation conducted through a coin toss | ✕ Researchers emailed participants to inform them which group they were assigned to | ✓ | ✕ Unable to blind due to nature of intervention | ✕ Unable to blind due to nature of intervention | ✓ | ✕ No mention | ✓ | ✓ Survey | ✓ 75% of participants completed all surveys. Differences btwn groups analysed and described | ✓ ITT analysis conducted | ✓ | ✓ |
| Ek, 2020 | ✓ Computer-generated random allocation sequence list generated by the study statistician | ✓ Allocation concealment was ascertained through opaque envelopes | ✓ | ✕ Group allocation was not blinded to the participants, who received an email after randomization | ✕ No mention | ✓ | ✓ | ✓ | ✓ Accelerometer, app and survey | ✓ LTFU adequately described. Sensitivity and attrition analysis conducted | ✓ ITT analysis conducted | ✓ | ✓ |
| Fyhri, 2015 | ✕ Not adequately described | ✕ No mention | ✕ Similar age, gender and employment btwn groups at baseline. Difference in cycling at baseline | ✕ Unable to blind due to nature of intervention | ✕ No mention | ✓ | ✕ No mention | ✓ | ✓ Survey, diary and odometer | ✓ | ✕ ITT analysis not reported | ✓ | ✓ Field exp |
| Grimes, 2020 | ✓ Random number generator used | ✓ | ✕ Sig diff in distance lives from campus | ✕ No mention | ✕ No mention | ✓ | ✕ No mention | ✓ | ✓ Activity tracker watch and pre/post survey | ✕ Not reported | ✕ ITT analysis not reported | ✓ | ✓ |
| Isensee, 2018 | ✓ Randomisation performed with a computer program | ✓ | ✕ Sig diff in type of school and days with at least 60mins of MVPA | ✕ No mention | ✕ No mention | ✓ | ✕ No mention | ✓ | ✓ Survey, self-reported | ✓ Attrition analysis | ✓ ITT analysis conducted | ✓ | ✓ Cluster RCT. Intra-cluster correlation reported. |
| King, 2017 | ✓ Random assignment using a web-based system | ✓ | ✓ | ✓ Single-blinded trial | ✕ Not blinded | ✓ | ✕ Not blinded | ✓ | ✓ Survey | ✕ High response rate but LTFU not adequately described | ✓ ITT analysis conducted | ✓ Mixed-effects regression models | ✓ |
| Liddle, 2014 | ✓ Random allocation via a computer-generated sequence concealed in opaque envelopes | ✓ | ✓ | ✕ Unable to blind due to nature of intervention | ✕ Not blinded | ✓ | ✓ | ✓ | ✓ Self-reported outcomes: home visit, telephone or mailed survey | ✓ LTFU adequately described. Attrition analysis conducted | ✓ ITT analysis conducted | ✓ | ✓ |
| Maca, 2020 | ✕ Not adequately described | ✕ No mention | ✕ Baseline demographics not reported adequately. | ✕ No mention | ✕ No mention | ✓ | ✕ No mention | ✓ | ✓ Survey | ✕ High dropout. Not adequately reported | ✕ ITT analysis not reported | ✓ Negative binomial regression model | ✓ |
| Mackey, 2019 | ✓ Web-based randomisation tool used | ✕ Study coordinator informed all participants of their assignments | ✓ | ✕ No mention | ✕ No mention | ✓ | ✓ Group assignment was concealed to outcome assesors | ✓ | ✓ Survey, travel diary and accelerometer | ✓ LTFU adequately described. | ✓ ITT analysis conducted | ✓ | ✓ |
| Mendoza, 2017 | ✓ Matched pairs. Random number assignment by study statistician | ✕ No mention | ✕ Difference in gender | ✕ Unable to blind due to nature of intervention | ✕ Unable to blind due to nature of intervention | ✓ | ✕ Unable to blind due to nature of intervention | ✓ | ✓ Survey, accelerometry and GPS units | ✓ No dropout | ✓ ITT analysis conducted | ✓ Repeated measures linear mixed effects model | ✓ Cluster RCT. Intra-cluster correlation reported. |
| Mendoza, 2011 | ✓ Matched randomisation. Random assignment by drawing the study condition from a container | ✕ No mention | ✕ Sig diff in distnace to school and active commuting at baseline | ✕ Unable to blind due to nature of intervention | ✕ Unable to blind due to nature of intervention | ✓ | ✕ Unable to blind due to nature of intervention | ✓ | ✓ Survey and accelerometer | ✓ LTFU adequately described, high response rate | ✓ ITT analysis conducted | ✓ | ✓ Pilot cluster RCT. Intra-cluster correlation reported. |
| Merom, 2007 | ✓ Computer-generated random numbers, stratified by gender were used to randomise participants | ✓ No mention, assumed it was done | ✓ | ✕ No mention | ✕ No mention | ✓ | ✓ | ✓ | ✓ Telephone interview | ✓ High response rate with no differences among groups. Adequately reported | ✓ ITT analysis conducted | ✓ | ✓ |
| Mutrie, 2002 | ✓ Matched respondents on distance travelled to work. Data management system randomly assigned group | ✓ | ✕ Demographics not adequately reported | ✕ Unable to blind due to nature of intervention | ✕ No mention | ✓ | ✕ | ✓ | ✓ Survey | ✓ LTFU reported, reasons provided. | ✓ ITT analysis conducted | ✓ | ✓ Stepwise logistic regression |
| Purath, 2005 | ✕ Not adequately described | ✕ No mention | ✕ Demographics not adequately reported | ✕ No mention | ✕ No mention | ✓ | ✕ No mention | ✓ | ✓ Survey | ✕ Not reported | ✕ ITT analysis not reported | ✓ | ✓ |
| Purath, 2004 | ✕ Not adequately described | ✕ No mention | ✕ Sig diff in race | ✕ No mention | ✕ No mention | ✓ | ✕ No mention | ✓ | ✓ Survey | ✓ LTFU adequately described, attrition analysis | ✕ ITT analysis not reported | ✓ | ✓ |
| Rowland, 2003 | ✓ An independent statistician carried out the randomisation using the MINIM software programme. | ✓ | ✓ | ✕ No mention | ✕ No mention | ✓ | ✕ No mention | ✓ | ✓ Survey | ✓ LTFU reported, reasons provided. | ✓ ITT analysis conducted | ✓ | ✕ Cluster RCT. Intra-cluster correlation not reported. |
| Salinas, 2019 | ✕ Simple randomisation conducted. Don't think this counts as true randomisation. | ✕ Study staff knew which community centres were in the intervention or control conditions | ✕ Sig diff in BMI | ✓ While study staff knew which community centres were in the intervention or control conditions, participants were unaware of what the status was of other centres. | ✕ Not blinded | ✓ | ✕ Not blinded | ✓ | ✓ Accelerometer and interview | ✕ Not adequately reported | ✓ ITT analysis conducted | ✓ | ✓ |
| Simon, 2014 | ✕ Not adequately described | ✕ No mention | ✕ Sig diff | ✕ No mention | ✕ Not blinded | ✓ | ✕ Not blinded | ✓ | ✓ Survey | ✓ LTFU adequately described. | ✓ ITT analysis provided in rationale | ✓ | ✓ Cluster RCT. Intra-cluster correlation reported. |
| Simon, 2008 | ✕ Not adequately described | ✕ No mention | ✕ Sig diff | ✕ No mention | ✕ Not blinded | ✓ | ✕ Not blinded | ✓ | ✓ Survey | ✓ LTFU adequately described | ✓ ITT analysis conducted | ✓ | ✓ Cluster RCT. Intra-cluster correlation reported. |
| Singh, 2009 | ✓ Randomisation was stratified. Randomisation using SPSS statistical software | ✕ No mention | ✕ Sig diff | ✕ No mention | ✕ Not blinded | ✓ | ✕ Not blinded | ✓ | ✓ Survey | ✓ LTFU adequately described | ✓ ITT analysis conducted | ✓ | ✓ |
| Sirard, 2008 | ✕ Not adequately described | ✕ Families were notified of their group randomisation | ✕ Diff in gender? | ✕ Not blinded | ✕ Not blinded | ✓ | ✕ No mention | ✓ | ✓ Accelerometer | ✓ LTFU adequately described | ✕ ITT analysis not reported | ✓ | ✓ |
| Suchert, 2015 | ✓ Stratified randomisation carried out using computer program | ✕ No mention | ✕ Sig diff | ✕ No mention | ✕ No mention | ✓ | ✕ No mention | ✓ | ✓ Survey | ✓ LTFU adequately described, attrition analysis | ✓ ITT analysis conducted | ✓ | ✓ Cluster RCT. Intra-cluster correlation reported. |
| Van Dyck, 2016 | ✓ Random selection and allocation using a computerised random number generator. | ✓ | ✕ Sig diff in age and gender | ✕ No mention | ✕ No mention | ✓ | ✕ No mention | ✓ | ✓ Survey | ✓ Dropout analysis | ✕ ITT analysis not conducted | ✓ | ✓ |
| van Stralen, 2010 | ✕ Not adequately described | ✕ No mention | ✕ Sig diff in education, BMI and self-efficacy | ✕ No mention | ✕ No mention | ✓ | ✕ No mention | ✓ | ✓ Survey | ✓ Dropout analysis | ✕ ITT analysis not reported | ✓ | ✕ Cluster RCT. Intra-cluster correlation not reported. |
| Xu, 2015 | ✓ Random allocation via random number generator software | ✓ | ✓ | ✕ No mention | ✕ No mention | ✓ | ✕ No mention | ✓ | ✓ Survey | ✓ LTFU adequately described | ✕ ITT analysis not reported | ✓ | ✕ Cluster RCT. Intra-cluster correlation not reported. |

Table 4. Extended quality assessment of quasi-experimental studies

| First author, Year of publication | 1. Is it clear in the study what is the ‘cause’ and what is the ‘effect’ (i.e. there is no confusion about which variable comes first)? | 2. Were the participants included in any comparisons similar? | 3. Were the participants included in any comparisons receiving similar treatment/care, other than the exposure or intervention of interest? | 4. Was there a control group? | 5. Were there multiple measurements of the outcome both pre and post the intervention/exposure? | 6. Was follow up complete and if not, were differences between groups in terms of their follow up adequately described and analysed? | 7. Were the outcomes of participants included in any comparisons measured in the same way? | 8. Were outcomes measured in a reliable way? | 9. Was appropriate statistical analysis used? |
| --- | --- | --- | --- | --- | --- | --- | --- | --- | --- |
| Aldred, 2021 | ✓ | ✓ | ✓ | ✓ | ✓ | ✕ | ✓ | ✓ | ✕ |
| Aldred, 2019 | ✓ | ✓ | ✓ | ✓ | ✕ | ✕ | ✓ | ✓ | ✕ |
| Auchincloss, 2019 | ✓ | ✓ | ✓ | ✓ | ✕ Pre/post only | ✕ Not reported | ✓ | ✓ | ✓ |
| Benton, 2021 | ✓ | ✓ | ✓ | ✓ | ✓ Observations at baseline and 7, 12 and 24 months post-baseline | ✕ Not reported | ✓ | ✓ | ✓ |
| Biondi, 2022 | ✓ | ✕ Demographics not provided | ✓ | ✓ | ✓ | ✕ Not reported | ✓ | ✓ | ✓ |
| Boarnet, 2005 | ✓ | ✕ Differences in income, race and neighbourhood form | ✕ | ✓ Children who didn't pass intervention were considered control/comparison | ✕ Pre/post only | ✕ 69% response overall, difference between the groups not adequately reported | ✓ | ✓ Survey only | ✓ |
| Brown, 2016 | ✓ | ✕ Significant differences in demographics. Near (intervention) residents were significantly less likely to report being white, married, or employed. | ✓ | ✓ | ✕ Measurement at Time 1 and Time 2 | ✕ Not adequately reported | ✓ | ✓ | ✓ Sample size met power req |
| Brown, 2003 | ✓ | ✕ Demographics not provided | ✓ | ✓ | ✕ Measurement before intervention and during intervention (after it had operated for 6m) | ✕ Not reported | ✓ | ✓ Survey only | X No mention of statistical procedure |
| Brownson, 2004 | ✓ | ✓ | ✓ | ✓ | X Pre/post | ✕ Not adequately reported | ✓ | ✓ Survey only | ✓ |
| Buckley, 2013 | ✓ | ✕ Demographics not provided | ✓ | ✓ | ✓ Fall study: pre, during and post. Spring study: x2 pre, during and x2 post | ✕ Not possible to determine a response rate | ✓ | ✓ Count data + survey | X No mention of statistical procedure |
| Bungum, 2014 | ✓ | ✓ Researchers matched intervention and control schools on size, demographics (SES and ethnicity) and built environment. No baseline demographics table. | ✓ | ✓ | ✕ Pre, during and post measurements | ✕ Not adequately reported | ✓ | ✓ | ✓ Chi-square test seems appropriate. No mention of power. |
| Cambra, 2020 | ✓ | ✕ Demographics not provided for both groups | ✓ | ✓ | ✕ Pre/post only | ✕ Not reported | ✓ | ✓ Pedestrian counts | ✓ Paired t-test used to compare baseline and follow-up data |
| Clark, 2014 | ✓ | ✓ Comparison trails matched neighborhood demographics as closely as possible | ✓ | ✓ | ✕ Pre, during and post measurements | ✕ Not possible to determine a response rate | ✓ | ✓ Infrared sensor and manual count audits | ✓ Non-parametric tests used |
| Coombes, 2016 | ✓ | ✓ No statistically significant differences between the groups in terms of gender and age | ✓ | ✓ | ✕ Pre, during and post measurements | ✓ Participant dropout adequately described; ITT analysis conducted | ✓ | ✓ Accelerometer and travel diary | ✓ |
| Crane, 2017 | ✓ | ✕ Significant differences in demographics. | ✓ | ✓ | ✓ Baseline and x2 follow-up | ✕ Not adequately reported | ✓ | ✓ Travel diary and survey | ✓ |
| Crawford, 2013 | ✓ | ✕ Sig diff for Education level | ✓ | ✓ Phase 1 controlled | ✕ Pre/post only | ✕ Not reported | ✓ | ✓ Bicycle counts and classroom survey | ✓ |
| Dill, 2014 | ✓ | ✓ | ✓ | ✓ | ✕ Pre/post only | ✕ Varied retention rate, higher in intervention area. Lack of description for LTFU reasons | ✓ | ✓ GPS, accelerometer and survey | ✓ |
| Elinder, 2012 | ✓ | ✓ | ✓ | ✓ | ✓ | ✓ Dropout analysis conducted | ✓ | ✓ Survey only for outcomes of interest. Diary and accelerometer used to test validity only. | ✓ |
| Felix, 2020 | ✓ | ✕ Demographics not adequately reported | ✓ | ✓ | ✓ | ✕ Not reported | ✓ | ✓ Bicycle counts | ✓ |
| Fitzhugh, 2010 | ✓ | ✕ Demographics not adequately reported | ✓ | ✓ | ✕ Pre/post only | ✕ Not reported | ✓ | ✓ PA and AT counts | ✓ |
| Folta, 2013 | ✓ | ✕ Significant diff in race/ethnicity | ✓ | ✓ | ✕ Pre/post only | ✓ Lower response rate at post, analysis conducted to account for missing data | ✓ | ✓ Survey only | ✓ |
| Frank, 2021 | ✓ | ✕ Significant diff for ethnicity | ✓ | ✓ | ✕ Pre/post only | ✕ Low response rate, 47% attrition rate. LTFU not reported adequately | ✓ | ✓ Survey and travel diary | ✓ Two tailed t-tests |
| Fuller, 2012 | ✓ | ✕ Demographics not reported. | ✓ | ✓ | ✓ | ✕ Not reported | ✓ | ✓ Objective bikeshare data | ✓ Interrupted time series approach + Bayesian structural time series model |
| Fuller, 2019 | ✓ | ✕ ITS, no comparison group | ✕ ITS, no comparison group | ✕ ITS, no comparison group | ✓ | ✕ Not reported | ✓ | ✓ Objective bikeshare data | ✓ Separate segmented regression models and Durbin-Watson stats |
| Geng, 2020 | ✓ | ✓ No statistically significant differences between groups | ✓ | ✓ | ✕ Pre/post only | ✕ Not reported | ✓ | ✓ Survey only | ✓ |
| Goodman, 2013 | ✓ | ✓ Intervention towns were similar to the matched comparison group in terms of population size, population density and affluence, and were also reasonably similar to the national comparison group | ✓ | ✓ | ✕ Pre/post only | ✕ Not adequately reported | ✓ | ✓ | ✓ Meta-regression |
| Goodman, 2016 | ✓ | ✕ Sig diff | ✓ | ✓ | ✕ Pre/post only | ✓ Analysis adjusted for missing data | ✓ | ✓ Survey only | ✓ Regression analyses |
| Grunseit, 2019 | ✓ | ✕ ITS, no comparison group | ✕ ITS, no comparison group | ✕ ITS, no comparison group | ✓ | ✕ Not reported | ✓ | ✓ Bicycle and pedestrian counts | ✓ Dickey-Fuller and Phillips-Perron unit root tests. ARIMA models. |
| Gu, 2019 | ✓ | ✕ Demographics not reported | ✓ | ✓ | ✕ Pre/post | ✕ Not reported | ✓ | ✓ Bicycle travel data | ✓ |
| Gutierrez, 2014 | ✓ | ✓ Control group schools were matched within-group intervention schools based upon selected demographic and injury rate criteria | ✓ | ✓ | ✓ x2 Pre and x2 post | ✕ Not adequately reported | ✓ | ✓ Bicycle and pedestrian counts, survey | ✓ ANOVA |
| He, 2022 | ✓ | ✓ Control groups consisted of participants residing in station catchments that were comparable in neighbourhood types, regional accessibility, socioeconomic status and demographics | ✓ | ✓ | ✕ Pre/post only | ✕ Low response rate | ✓ | ✓ Survey only for outcomes of interest. | ✓ DID regression analyses |
| Heelan, 2009 | ✓ | ✓ | ✓ | ✓ | ✓ 6 measurement times | ✕ LTFU not adequately reported. Missing data removed from analyses | ✓ | ✓ Survey and accelerometer | ✓ Repeated measures ANOVA |
| Heesch, 2016 | ✓ | ✕ Differences in gender. | ✓ | ✓ "Controls” were the SEFB and Logan Road | ✕ Pre/post only | ✓ Response rate reported. No significant differences in gender or riding composition between cyclists who completed the survey and those who did not, for any survey (p>0.05). | ✓ | ✓ Field observations, GPS bicycle count data and intercept survey | ✓ |
| Heinrich, 2011 | ✓ | ✕ Sig diff | ✓ | ✓ | ✕ Pre/post only | ✕ Variable response rates amongst the different schools. Lack of adequate description or analysis. | ✓ | ✓ Parent survey, classroom travel, traffic counts and safety observations | ✕ Statistical analysis methods not described in detail |
| Hoelscher, 2016 | ✓ | ✓ | ✓ | ✓ | ✕ Pre/post only | ✕ Not adequately reported | ✓ | ✓ Self-reported only. Student and parent surveys | ✓ Mixed linear regression and growth curve models |
| Hong, 2016 | ✓ | ✓ No statistically significant differences between treatment and control group | ✓ | ✓ | ✕ Pre/post only | ✓ Response rate reported and did not vary greatly by household and demographic characteristics. | ✓ | ✓ Accelerometer, GPS device and survey | ✓ |
| Hosford, 2019 | ✓ | ✓ | ✓ | ✓ | ✓ Pre, 2x post | ✕ Not adequately reported | ✓ | ✓ Survey | ✓ Regression analysis and modelling |
| Hunter, 2021 | ✓ | ✕ Demographics not adequately reported | ✓ | ✓ | ✕ Pre/post | ✓ LTFU adequately described. | ✓ | ✓ Survey | ✓ |
| Jancey, 2011 | ✓ | ✓ No statistically significant differences between groups | ✓ | ✓ | ✓ Baseline, postintervention | ✓ Similar dropout rate for both groups. Demographic characteristics of dropouts were analysed, no significant differences. | ✓ | ✓ Survey | ✓ |
| Jones, 2012 | ✓ | ✓ Both neighbourhoods reported as being demographically similar | ✓ | ✓ | ✕ Post only | ✕ Not reported | ✓ | ✓ Survey | ✓ |
| Jordan, 2008 | ✓ | ✓ | ✓ | ✓ | ✕ Pre/post only | ✕ LTFU not adequately reported. | ✓ | ✓ Survey | ✓ |
| Karpinski, 2021 | ✓ | ✕ Demographics not reported | ✓ | ✓ | ✓ | ✕ Not reported | ✓ | ✓ Trip data from bikeshare program | ✓ Analysis of variance |
|  |  |  |  |  |  |  |  |  | and Spearman and Pearson correlations were used |
| Keall, 2022 | ✓ | ✓ Control cities chosen due to similar geography, demographics, economic profiles and climated to intervention cities | ✓ | ✓ | ✓ 5 measurements (pre and 4x subsequently) | ✕ Not reported | ✓ | ✓ Survey | X No mention of statistical procedure |
| Keall, 2015 | ✓ | ✓ Control cities have similar demographics, economic profiles and climates to intervention cities | ✓ | ✓ | ✕ Pre, during & post | ✕ Variable response rates. Lack of adequate description or analysis. | ✓ | ✓ Survey | ✓ Generalised linear mixed models |
| Kubota, 2020 | ✓ | ✕ Sig diff for Education level | ✓ | ✓ | ✕ Pre/post | ✓ Similar response rate at both time points. Missing data excluded from analysis; demographics described for participants removed | ✓ | ✓ Survey | ✓ Generalised linear models |
| Lambe, 2022 | ✓ | ✕ Sig diff for gender | ✕ 26% of control group were aware of intervention. Seems like control schools were in same area as intervention schools and may have been exposed | ✓ | ✕ Pre/post | ✕ High response rate at both time points, however description and analysis not adequately reported. | ✓ | ✓ Survey | ✓ Chi-square test and binary logistic regression analysis |
| Lambe, 2017 | ✓ | ✕ Sig diff | ✓ | ✓ | ✕ Pre/post | ✕ Not adequately reported | ✓ | ✓ Survey | ✓ |
| Limb, 2020 | ✓ | ✕ Sig diff | ✓ | ✓ | ✕ Pre/post | ✕ Not adequately reported | ✓ | ✓ Accelerometry, GPS travel recorder | ✓ Multilevel linear regression models |
| Malakellis, 2017 | ✓ | ✕ Sig diff in BMI and parental education attainment | ✓ | ✓ | ✕ Pre/post | ✓ LTFU adequately described. | ✓ | ✓ Survey | ✓ |
| Mayer, 1982 | ✓ | ✕ Demographics not reported, unable to contact authors for data, | ✓ | ✓ | ✕ Pre/post | ✕ Not adequately reported | ✓ | ✓ Survey and frequency count | ✓ |
| McDonald, 2014 | ✓ | ✕ Sig diff | ✓ | ✓ | ✓ Pre, 6x post | ✕ Not reported | ✓ | ✓ Survey and interview | ✓ Modelling |
| McKee, 2007 | ✓ | ✕ Sig diff | ✓ | ✓ | ✕ Pre/post | ✕ High response rate at both time points, however description and analysis not adequately reported. | ✓ | ✓ Computerised mapping programme and survey | ✓ |
| McMinn, 2012 | ✓ | ✕ Difference in walkers | ✓ | ✓ | ✓ Data collected over 5 days, reported as pre/post | ✕ Not reported | ✓ | ✓ Accelerometer, survey and travel diary | ✓ ANOVA |
| Mendoza, 2009 | ✓ | ✓ | ✓ | ✓ | ✓ Pre, 3x post | ✕ Not adequately reported. Similar response rate at each time point | ✓ | ✓ Survey | ✓ |
| Merom, 2003 | ✓ | ✕ Sig diff in gender | ✓ | ✓ | ✕ Pre/post | ✓ LTFU adequately described. | ✓ | ✓ Survey and bike counts | ✓ |
| Mitra, 2021 | ✓ | ✓ | ✓ | ✓ | ✕ One measurement only? Retrospective survey design | ✕ Not reported | ✓ | ✓ Survey | ✓ |
| Olsen, 2016 | ✓ | ✓ | ✓ | ✓ | ✕ Pre/post | ✕ Not reported | ✓ | ✓ Travel diary and survey | ✓ Logistic regression model |
| Ostergaard, 2015 | ✓ | ✕ Sig diff | ✓ | ✓ | ✕ Pre/post | ✕ Not reported | ✓ | ✓ Survey | ✓ |
| Petrunoff, 2016 | ✓ | ✕ ITS, no control | ✕ ITS, no control | ✕ ITS, no control | ✓ 4 measurements | ✕ Not adequately reported | ✓ | ✓ Survey | ✓ |
| Rissel, 2010 | ✓ | ✓ | ✓ | ✓ | ✕ Pre/post | ✓ LTFU characteristics described | ✓ | ✓ Survey and bicycle counters | ✓ |
| Rissel, 2015 | ✓ | ✕ Demographics not adequately reported for both groups separately | ✓ | ✓ | ✕ Pre/post | ✓ LTFU adequately described. | ✓ | ✓ Survey and travel diary | ✓ |
| Rothman, 2022 | ✓ | ✓ Schools were matched at school level and SES | ✓ | ✓ | ✕ Pre/post | ✕ Not adequately reported. | ✓ | ✓ Manual observations of AST | ✓ |
| Sahlqvist, 2019 | ✓ | ✕ Sig diff | ✕ 'Non-participating children' could have still been exposed to intervention | ✓ Comparison group was 'non-participating children' | ✕ Pre, during & post | ✕ Not adequately reported. | ✓ | ✓ Survey | ✓ Generalised Linear Models |
| Sersli, 2019 | ✓ | ✕ Sig diff | ✓ | ✓ | ✓ Baseline, 1, 3, and 12 months post | ✕ High response rate, however description and analysis not adequately reported. | ✓ | ✓ Survey | ✓ |
| Spears, 2016 | ✓ | ✓ | ✓ | ✓ | ✓ Pre and 2x post | ✓ LTFU adequately described. | ✓ | ✓ Survey, travel log and odometer | ✓ DID regression analysis |
| Stappers, 2021 | ✓ | ✕ Sig diff in age | ✓ | ✓ | ✕ Pre/post | ✓ Dropout analysis conducted | ✓ | ✓ Accelerometer, GPS and survey | ✓ |
| Sun, 2020 | ✓ | ✕ Differences in car ownership and education | ✓ | ✓ | ✕ Pre/post | ✓ LTFU adequately described. | ✓ | ✓ Survey | ✓ |
| Sundfor, 2022 | ✓ | ✕ Sig diff in age | ✓ | ✓ | ✕ Pre/post | ✕ Not adequately reported. | ✓ | ✓ Survey including travel diary and mobile app | ✓ |
| Villa-Gonzalez, 2017 | ✓ | ✕ Sig diff for distance from home to school | ✓ | ✓ | ✕ Pre/post | ✕ Not adequately reported. | ✓ | ✓ Survey | ✓ |
| Villa-Gonzalez, 2016 | ✓ | ✓ | ✓ | ✓ | ✓ Pre, post, 6m follow up | ✕ High dropout, reasons not reported | ✓ | ✓ Survey | ✓ Kolmogorov-Smirnov test? |
| Walsh, 2021 | ✓ | ✕ Sig diff | ✓ | ✓ | ✕ Pre/post | ✓ Low attrition rate, reasons for LTFU provided | ✓ | ✓ Survey and travel diary | ✓ |
| Xiao, 2022 | ✓ | ✕ Demographics not reported | ✕ | ✓ | ✓ | ✕ Not reported | ✓ | ✓ Cycle count data | ✓ ITS |
