## Supplementary material for "Effectiveness of interventions for modal shift to walking and bike riding: a systematic review with meta-analysis": Data Supplement 5

Data Supplement 5 – Results Tables

Table 1. Effect of interventions in cycling duration

| **First author, Year of publication** | **Intervention summary** | **Measure description** | **Intervention mean change (SD)** | **Control mean change (SD)** | **Mean difference** | **95% CI** |
| --- | --- | --- | --- | --- | --- | --- |
| Aittasalo, 2012 & 2019 | Workplace-specific educational programme | Minutes bicycled part of the journey from work | 2.5 (14.3) | 1.1 (12.0) | 1.4 | (-0.7 - 3.5) |
| Geng, 2020* | General health messaging | Minutes spent per day travelling by bicycle in the last week | 0.3 | -0.05 | 0.4 | Not reported |
| Salinas, 2019* | Intensive educational programme | Minutes cycling per week, mean (SD) | 25.0 (7.4) | 11.0 (3.9) | 14.0 | (8.8 – 19.2) |
| Van Dyck, 2016* | Individually tailored behavioural programme | Minutes per week cycling for transport, mean (SD) | 6.1 (92.9) | -0.4 (103.4) | 6.5 | (-16.5 - 29.5) |
| Van Stralen, 2010** | Individually tailored behavioural programme | Minutes per week cycling for transport, mean (SD) | Basic: 3.1 (79.5) Tailored: 5.1 (92.1) | 0.3 (67.9) | Basic: 2.8  Tailored: 4.8 | Basic: (-5.4 - 11.0) Tailored: (-3.8 - 13.4) |
| Ducheyne, 2014 | School based cycle training | Time spent cycling to school, minutes weekly | -4.1 (24.6) | 3.5 (30.4) | -7.6 | (-21.5 - 6.3) |
| Crane, 2017* | Supportive active transport infrastructure (e.g. cycleways and traffic calming) | Cycling minutes per week, mean (SD) | 3.0 (152.4) | -25.0 (125.6) | 28.0 | (9.0 – 47.0) |
| Gu, 2019 | New public transport service (e.g. new train line or light rail service) | Average cycling trip time, minutes | 1.1 | -0.4 | 1.5 | Not reported |
| Limb, 2020 | Supportive active transport infrastructure (e.g. cycleways and traffic calming) | Minutes cycling per week, mean (SD) | 30 (47) | 1 (92) | 29 | (17 – 41) |
| Rissel, 2010 | Community based cycling promotion program | Minutes cycling for travel | 115.0 | -78.3 | 193.3 | Not reported |

**Included in meta-analysis, **Tailored intervention included in meta-analysis*

Table 2. Effect of interventions in cycling frequency (proportions)

| **First author, Year of publication** | **Intervention summary** | **Measure description** | **Intervention mean change, %** | **Control mean change, %** | **Mean difference %** | **95% CI** |
| --- | --- | --- | --- | --- | --- | --- |
| Lambe, 2022 | Gamification of walking to school | Active travel from school percentage cycling | -0.8 | 3.4 | -4.2 | (-5.5 - 1.5) |
| Crane, 2017 | Supportive active transport infrastructure (e.g. cycleways and traffic calming) | Percentage cycling at least weekly | 6.0 | 3.6 | 2.4 | (-0.1 – 3.1) |
| Goodman, 2013 | Supportive active transport infrastructure (e.g. cycleways and traffic calming) | Prevalence of cycling to work (%) | 0.9 | 0.3 | 0.6 | Not reported |
| Jones, 2012 | Supportive active transport infrastructure (e.g. cycleways and traffic calming) | Proportion of trips made by bike (%) | -1 | 0 | -1 | (-2.4 - 2.1) |
| Bungum, 2014 | Designated ride and walk to school days | Proportion of students who cycling to school (%) | 0.1 | 0.1 | 0.0 | (-0.9 - 1.2) |
| Rissel, 2010 | Community based cycling promotion program | Proportion who have cycled in last year (%) | -0.6 | 3.4 | -4.0 | Not reported |
| Bjornara, 2019 | Provision of bikes or e-bikes | Proportion of people cycling to work (%) | 38.9 | 0.3 | 38.6 | Not reported |
| Mendoza, 2017 | School based bicycle train | Proportion who cycle daily to school (%) | 54.1 | 12.0 | 42.1 | Not reported |

Table 3. Effect of interventions on cycling frequency (means)

| **First author, Year of publication** | **Intervention summary** | **Measure description** | **Intervention mean change (SD)** | **Control mean change (SD)** | **Mean Difference** | **95% CI** |
| --- | --- | --- | --- | --- | --- | --- |
| Villa-Gonzalez, 2016 & 2017 | School based educational program | Cycling trips per week, mean (SD) | 0.0 (0.0) | -0.3 (0.2) | 0.3 | Not reported |
| Sundfor, 2022 | Financial subsidy for e-bike purchase | Number of bicycle trips over study period (two months), mean (SD) | 0.1 (0.8) | 0.4 (1.2) | 0.3 | (0.2 – 0.4) |
| Aittasalo, 2019 | Workplace-specific educational programme | Number of days bicycling from work per week, mean (SD) | 0.3 (1.7) | 0.1 (1.9) | 0.2 | (-0.1 - 0.5) |
| Sersli, 2019 | Bicycle skills training | Bicycling frequency days/month mean (SD) | 0.8 (12.3) | -1.3 (13.2) | 2.1 | (-9.9 – 14.1) |
| Frank, 2021 | Supportive active transport infrastructure (e.g. cycleways and traffic calming) | Cycling trip count, mean | 0.10 | 0.03 | 0.07 | Not reported |
| Spears, 2016 | New public transport service (e.g. new train line or light rail service) | Mean number of bicycle trips per week | 0.1 | 0.1 | 0.0 | Not reported |
| Andersson, 2021 | Provision of e-bikes | Average number of trips travelled per day by bicycle, mean (SD) | 2.1 (4.2) | 0.7 (2.1) | 2.8 | (0.7 - 3.5) |
| Bernstein, 2017 | Provision of a bicycle with training materials | Used a bicycle 2+ times for of times in to shop/eat/visit friends or any other activity besides working the past 7 days (y/n) | 8.3 | -10.0 | 18.3 | Not reported |

**Difference in medians*

Table 4. Effect of interventions on walking duration

| **First author, Year of publication** | **Intervention summary** | **Measure description** | **Intervention mean change (SD)** | **Control mean change (SD)** | **Mean difference** | **95% CI** |
| --- | --- | --- | --- | --- | --- | --- |
| Aittasalo, 2019* | Workplace-specific educational programme | Minutes walked for part of the journey from work, mean (SD) | 0.3 (10.7) | -0.9 (12.1) | 1.2 | (-0.7 - 3.1) |
| Salinas, 2019 | Intensive educational programme | Minutes walking per week, mean difference (SD) | 82.0 (24.3) | 43.0 (15.2) | 39.0 | (21.1-46.3) |
| Aittasalo, 2012* | Workplace-specific educational programme | Weekly minutes walked, mean (SD) | 55.0 (259.1) | -7.0 (170.0) | 62.0 | (6.9 - 117.1) |
| Dunton, 2008* | Individually tailored behavioural programme | Weekly minutes walked, mean (SD) | 68.6 (207.7) | 32.4 (227.1) | 36.2 | (-38.5 - 110.9) |
| Merom, 2003 | Promotion of new walking infrastructure | Weekly walking hours, mean (SD) | 0.0 (3.3) | 0.1 (3.4) | 0.1 | (-0.7 - 0.7) |
| Merom, 2007 | Individually tailored behavioural programme | Weekly minutes walked, mean difference (95% CI) | Not reported | Not reported | 2.2 | Not reported |
| Mutrie, 2002 | Individually tailored behavioural programme | Minutes walked to work per week, mean | 27 | 10 | 17 | Not reported |
| Purath, 2004* | Individually tailored behavioural programme | Weekly minutes walked, mean (SD) | 103.1 | 76.2 | 26.9 | Not reported |
| Van Dyck, 2016* | Individually tailored behavioural programme | Weekly minutes walked for transport, mean (SD) | 24.7 (134.7) | -0.2 (112.6) | 24.9 | (-4.8 - 54.6) |
| Van Stralen, 2010 | Individually tailored behavioural programme | Weekly minutes walked for transport, mean (SD) | Group 1: 3.4 (47.8) Group 2: 1.1 (45.3) | Group 1: -1.2 (50.1) Group 2: 605.2 (423.8) | Group 1: 4.6 Group 2: -604.1 | Group 1: (-0.9 - 10.1) Group 2: (-638.6 - -569.6) |
| King, 2017^(76)^ | Individually tailored behavioural programme | Weekly minutes walked for errands, mean | 32.6 | 48.0 | -15.4 | Not reported |
| He, 2022* | New public transport service (e.g. new train line or light rail service) | Weekly minutes walked, mean (SD) | 59.8 (463.8) | 10.9 (447.7) | 48.9 | (-10.7 - 108.5) |
| Limb, 2020* | Supportive active transport infrastructure (e.g. cycleways and traffic calming) | Weekly minutes walked, mean (SD) | -0.7 (23.8) | -0.7 (25.5) | 0 | (-4.0 - 4.0) |
| Jancey, 2011 | Walking program with personal advice call centre for adults | Weekly minutes walked, mean (SE) | 155.8 (8.7) | 26.2 (8.9) | 129.5 | (121.1 – 137.2) |
| Kubota, 2020 | Provision of walking maps and events | Daily minutes walked, mean (SD) | -16.2 (87.3) | 3.2 (95.8) | -19.4 | (-33.6 - -5.2) |

**Included in meta-analysis*

Table 5. Effect of interventions on walking participation (proportions)

| **First author, Year of publication** | **Intervention summary** | **Measure description** | **Intervention mean change (%)** | **Control mean change (%)** | **Effect estimate (%)** | **95% CI** |
| --- | --- | --- | --- | --- | --- | --- |
| Geng, 2020 | General health messaging | Frequency of two-way trips by foot in the last week | Intervention 2: -0.50 | -0.06 | Intervention 2: -0.44 | Not reported |
| Aittasalo, 2012 | Workplace-specific educational programme | Frequency of walkers for transport purposes | 7.3 | 6.7 | 0.6 | (-0.2 – 2.6) |
| Mackey, 2019 | Individually tailored behavioural programme | Frequency of walking trips number per day | -0.2 | -0.3 | 0.1 | (-3.3 - 3.4) |
| Lambe, 2022 | Gamification of walking to school | Proportion of trips to school by walking (%) | 0.5 | -4.7 | 5.3 | (-17.8 – 8.4) |
| Bungum, 2014* | Designated ride and walk to school days | Mode of transport to school is walking | -8 | -8 | 0 | (-1 - 1) |
| Mendoza, 2009 | School based walking bus | Frequency of transport to school by walking | 5 | -8 | 13 | Not reported |
| Rowland, 2003 | School based walking bus | Frequency of students who walked to school | 9.8 | 10.5 | -0.7 | Not reported |
| Xu, 2015 | School based educational campaign | Proportion of people who indicated they had increased walking frequency (%) | 46.0 | 32.4 | 13.6 | Not reported |
| Brown, 2003 | Fare-free public transport | Frequency of walking as the chosen commute mode for faculty & staff (%) | -98 | 11 | -109 | Not reported |

Table 6. Effect of interventions on walking participation (means)

| **First author, Year of publication** | **Intervention summary** | **Measure description** | **Intervention mean change (SD)** | **Control mean change (SD)** | **Mean difference** | **95% CI** |
| --- | --- | --- | --- | --- | --- | --- |
| Aittasalo, 2019 | Workplace-specific educational programme | Number of days walked for part of the journey from work, mean (SD) | 0.1 (1.9) | -0.3 (2.0) | 0.4 | (0.1 - 0.7) |
| Villa-Gonzalez, 2017 | School based educational program | Boys frequency of commuting to school by walking, per week, mean (SD)  Girls frequency of commuting to school by walking, per week, mean (SD) | -0.3 (0.4)  -0.2 (0.4) | 0.9 (0.6)  0.2 (0.6) | -1.1  -0.4 | Not reported |
| Villa-Gonzalez, 2016 | School based educational program | Frequency of commuting to school by walking, per week, mean (SE) | 1.1 (0.3) | -0.1 (0.4) | 1.2 | Not reported |
| Sundfor, 2022 | Financial subsidy for e-bike purchase | Frequency of walking trips, mean (SD) | -0.3 (1.1) | -0.1 (1.2) | -0.2 | (-0.3 - -0.1) |
| Spears, 2016 | New public transport service (e.g. new train line or light rail service) | Walk trips, mean | 0 (0) | 0 (0) | 0 | Not reported |
| Folta, 2013 | School based walking bus | Frequency of trips weekly to/from school, mean (SD) | 0.4 (4.2) | -0.1 (3.9) | 0.5 | (-0.4 - 1.4) |
| Kubota, 2020 | Provision of walking maps and events | Frequency of walking per week, mean (SD) | -0.1 (2.4) | 0 (2.4) | -0.1 | (-0.5 - 0.3) |
| Andersson, 2021 | Provision of e-bikes | Frequency of walking trips per day, mean (SD) | 0.7 | 0.2 (1.3) | 0.4 | Not reported |
